# Cost-effectiveness of subnational targeting of small-quantity lipid-based nutrient supplementation: A simulation model in Nigeria, Ethiopia, and Pakistan

**DOI:** 10.64898/2026.09.03.26362091

**Authors:** Alison Bowman, Sylvia Lutze, Zeb Burke-Conte, Nathaniel Deveroux Blair-Stahn, Alix Pletcher, Hussain Jafari, James R. Albright, Rajan Mudambi, Abraham D. Flaxman

## Abstract

**Introduction:** Small-quantity lipid-based nutrient supplementation (SQ-LNS) is an intervention that provides supplemental nutrition to young children and has significant impacts on mortality, nutrition, and developmental outcomes. The World Health Organization recommends the use of SQ-LNS in certain targeted contexts, but evidence on the cost-effectiveness of SQ-LNS targeting approaches is limited.

**Methods:** We utilized an individual-based simulation model to estimate the cost-effectiveness of SQ-LNS within first-level administrative divisions in Ethiopia, Nigeria, and Pakistan. We estimated the cost-effectiveness of subnational targeting strategies including threshold- and priority-based targeting algorithms with several indicators across child mortality, nutrition, and complementary feeding measures.

**Results:** Subnational targeting according to wasting, stunting, and anemia thresholds was on average more cost-effective than untargeted strategies for all locations. Incremental cost-effectiveness ratios for these targeted and untargeted strategies respectively were $363 (95% uncertainty interval, UI: 243, 666) per disability-adjusted life year (DALY) averted and $458 (95% UI: 291, 863) in Ethiopia, $135 (95% UI: 74.2, 305) and $227 (95% UI: 124, 513) in Nigeria, and $474 (95% UI: 329, 718) and $544 (95% UI: 366, 878) to in Pakistan. Priority-based targeting using a composite ranking measure across mortality, wasting, and stunting burdens offered the greatest cost-effectiveness of any strategy assessed, achieving 7.0% (UI: 4.4, 12) and 4.6% (UI: 1.4, 7.3) more DALYs averted for the same cost as the threshold-based targeting approach in Nigeria and Ethiopia and equal impact in Pakistan.

**Conclusions:** Subnational targeting of SQ-LNS can significantly improve cost-effectiveness relative to untargeted scale-up strategies. Several targeting strategies and indicators may be used to improve cost-effectiveness, with highest performance seen with a priority-based algorithm using a composite ranking measure across child mortality, wasting, and stunting burdens. Subnational targeting according to such measures should be considered in the implementation of SQ-LNS programs in accordance with existing guidelines.

## Introduction

There has been significant progress in addressing child undernutrition in recent years [1]. Even still, child growth failure (CGF), inclusive of child stunting, wasting, and underweight, remains a top contributor to global morbidity and mortality, estimated to cause over one million deaths in 2023 [1]. There is robust evidence that small-quantity lipid-based nutrient supplementation (SQ-LNS) provided daily to children six to 24 months of age is an effective intervention to prevent CGF. An individual participant data meta-analysis has shown that SQ-LNS causes significant improvements in child wasting and stunting in addition to mortality, anemia, and developmental outcomes [2]. Another recent analysis found that children who received SQ-LNS before receiving acute malnutrition (AM) treatment were less likely to be hospitalized for AM [3]. Initial analyses on the cost-effectiveness of SQ-LNS have found costs per disability-adjusted life year (DALY) averted to range from $242 in Uganda [4] to $747 in Mali and $1,073 in Burkina Faso [5]. While these analyses have varied conclusions on the cost-effectiveness classification of SQ-LNS, they each recognize that the total cost of scaling up a national SQ-LNS program would be high. To increase financial feasibility, strategies to reduce costs such as targeting coverage to populations of greatest need will likely be valuable.

While SQ-LNS has not yet been implemented at scale, there have been several efforts to identify specific criteria of populations that may benefit from SQ-LNS. The individual-participant data meta-analysis reported suggestive evidence that the effect of SQ-LNS on severe wasting was greater in populations without improved water quality and sanitation as well as in populations with higher baseline prevalence of stunting or wasting [6]. UNICEF’s guidance on SQ-LNS recommends it be used in nutritionally at-risk populations such as those with high stunting, wasting, and anemia prevalence with emphasis on those that also have high mortality, wasting treatment relapse rates, and micronutrient deficiencies [7]. Similarly, operational guidance prepared by the SQ-LNS Task Force outlines several specific measures and thresholds to consider in identifying appropriate populations for SQ-LNS programs, including wasting, stunting, anemia, and complementary feeding measures [8]. Noori et al. (2025) constructed quantitative national and subnational rankings for SQ-LNS prioritization based on a combination of wasting, stunting, and mortality measures [9].

In its 2023 guideline, the World Health Organization (WHO) endorsed the use of SQ-LNS for the prevention of wasting in contexts with high food insecurity through a targeted approach [10]. Despite moderate certainty around the effects of SQ-LNS, this recommendation was classified as low certainty overall due to reliance on indirect evidence to inform its use in contexts of high food insecurity and noted limited evidence on the cost-effectiveness of blanket versus targeted approaches.

To help address this gap, we extended a previously developed *in silico* microsimulation model [11] parameterized with 2021 Global Burden of Disease (GBD) estimates [12,13] to evaluate the performance of subnational SQ-LNS targeting in Ethiopia, Nigeria, and Pakistan. This analysis builds on the existing evidence and offers a quantitative comparison of the cost-effectiveness of multiple geographic targeting strategies for SQ-LNS programs, including threshold- and priority-based approaches informed from numerous measures related to child mortality, growth, and nutrition.

## Methods

### Health Impact Simulation

The health impact model used for this analysis was an individual-based simulation described in detail elsewhere [11]. In short, we initialized a cohort of live births and tracked outcomes through five years of life. We tracked CGF as four-category exposure measures for wasting (based on weight-for-height z-scores — WHZ), stunting (based on height-for-age z-scores —HAZ), and underweight (based on weight-for-age z-scores — WAZ). For each of the three measures, the exposure categories were severe (z-score < − 3), moderate (− 3 ≤ z-score < − 2), mild (− 2 ≤ z-score < − 1), and unaffected (z-score > − 1). We modeled wasting exposure with a dynamic transition model calibrated to GBD estimates at steady state. We assigned underweight exposure probabilistically based on current stunting and wasting exposures. In accordance with GBD 2021 risk effects models, we included causal pathways from child wasting, stunting, and underweight to the risk of morbidity and mortality due to diarrheal diseases, lower respiratory infections, malaria, and measles, as well as between child wasting and morbidity and mortality due to protein energy malnutrition [12]. We estimated DALYs in our simulation as the sum of years lived with disability (YLDs) due to modeled causes and years of life lost (YLLs) from overall mortality in accordance with GBD 2021 methodology [13].

Treatment for moderate and severe AM and SQ-LNS interventions were modeled as described in Bowman et al. (2025) [11]. AM treatment was included in this model as SQ-LNS is expected to reduce AM treatment costs by preventing wasting. Briefly, children with moderate or severe wasting between six and 59 months of age are eligible for AM treatment interventions that affect recovery rates. Simulated children covered by SQ-LNS began supplementation at six months and continued for a duration of 12 months. SQ-LNS affected wasting transition rates from less to more severe exposures as well as stunting exposure. In the present analysis, we performed a sensitivity analysis in which the effects of SQ-LNS were modified by subnational baseline under-five wasting prevalence above or below 10%. Effects of SQ-LNS on four-category wasting and stunting exposures for both the main and sensitivity analyses were informed from data obtained via correspondence with the authors of Dewey et al. (2021) [2]. Details of how they were applied are provided in Bowman et al. (2025) [11]. In the present analysis, we performed a post-hoc estimation of DALYs due to anemia averted by SQ-LNS in which we applied the effects of SQ-LNS on anemia from Wessells et al. (2021) [14] (total anemia: RR=0.84, 95% CI=0.81, 0.87; moderate to severe anemia: RR=0.72, 95% CI: 0.68, 0.76) to severity-specific anemia YLDs among the population six to 24 months of age as estimated by the GBD 2021 study.

We extended the health impact simulation described in Bowman et al. (2025) [11] to support first-level administrative division (Admin1) locations among our three modeled countries (Ethiopia, Nigeria, and Pakistan). We used GBD 2021 estimates specific to each Admin1 location to parameterize CGF exposures as well as disease incidence and mortality rates. We updated all modeled parameters that depend on these measures (such as wasting transition rates calibrated to wasting exposure and mortality rates) to be specific to the Admin1 location. All remaining parameters (such as birth weight and gestational age exposures and baseline intervention coverage estimates) were informed from national-level estimates.

Each Admin1 location was simulated with a population size of 400,000 pregnancies in Ethiopia and Pakistan and 200,000 pregnancies in Nigeria. We simulated 20 Monte Carlo uncertainty draws each in Ethiopia and Pakistan and 10 in Nigeria. We simulated a baseline scenario with 0% coverage of SQ-LNS and baseline coverage of moderate and severe AM treatment as reported in Bowman et al. (2025) [11].

We modeled an alternative scenario in which SQ-LNS was scaled-up to 70% coverage and coverage of AM treatment interventions remained at the baseline level. We assumed coverage of SQ-LNS and AM treatment interventions were independent at the individual level.

### Intervention Costing Approach

Our analysis included cost estimates from societal, governmental, and healthcare provider perspectives in alignment with available data. The cost of SQ-LNS was informed as an average of observed costs among studies included in the SQ-LNS Working Group’s technical brief on costs [15]. The costs for AM treatment were informed from Isanaka et al. (2019) [16] adjusted to 2021 USD, using ready-to-use-supplementary food as the moderate AM treatment product, and excluding management and administration costs as we assumed these would not scale directly with case load reductions on the scale caused by SQ-LNS. Modeled costs were $58.50 for 12 months of SQ-LNS, $38.82 per moderate AM case treated, and $134.68 per severe AM case in 2021 USD.

We made two updates to the SQ-LNS costing assumptions from the previously published methods [11]. First, we assumed that SQ-LNS costs did not accrue while a simulated child was on active treatment for AM. Second, we did not consider AM screening costs in this analysis.

### Targeting Indicators

The individual targeting indicators we considered were selected based on relevance to existing guidelines and literature [7–9] and included:

- Mortality risk (deaths per 1,000 live births),
- Wasting prevalence (WHZ < −2),
- Stunting prevalence (HAZ < −2),
- Underweight prevalence (WAZ < −2),
- Anemia prevalence (hemoglobin < 150 g/L among neonates and < 110 g/L otherwise), and
- Complementary feeding indicators, including prevalence of less than minimum:

- Dietary diversity
- Meal frequency
- Acceptable diet

All indicators were measured among the under-five population except the complementary feeding indicators that are specifically measured among children aged six to 24 months. We informed all indicators using GBD 2021 estimates except for the complementary feeding indicators, which we informed from the country-specific Demographic and Health Surveys from the available survey year closest to 2021 to maximize temporal consistency with GBD estimates used in our model [17–19]. We did not include a food insecurity indicator due to lack of publicly-available data at the Admin1 level.

The composite targeting indicators we considered included:

- Global Hunger Index (GHI) score, and
- Composite ranking across mortality, wasting, and stunting

We calculated GHI score as a weighted function of standardized under-five mortality risk, wasting and stunting prevalence, and underweight prevalence as a proxy for undernutrition according to published methodology [20] and used GBD 2021 estimates of underweight (WAZ < −2) prevalence as a proxy for undernourishment defined as insufficient caloric intake. We generated the composite ranking in the manner described by Noori et al. (2025) [9] as the overall rank across six to 24 month mortality rate (deaths per 1,000 live births), severe wasting (WHZ < −3) prevalence among the six to 24 month age group, and severe stunting (HAZ <-3) prevalence among the 24-60 month age group. We ordered composite rankings in ascending order such that locations with high ranks represent greater underlying burden.

### Analysis

At the scenario-, input draw-, and subnational location-specific level, we calculated total cost spent on each intervention by multiplying the scale of the intervention administered (cases treated for AM treatment and person-years covered by SQ-LNS) in the simulation by the assumed per unit intervention costs. We scaled the total costs and DALYs output from our simulation population to the expected number of live births in each subnational location according to GBD 2021 estimates. We calculated DALYs averted and incremental costs as the difference in the baseline at alternative scenarios. For the estimate of DALYs averted, we added in the post-hoc assessment of DALYs averted due to anemia. We calculated incremental cost-effectiveness ratios (ICERs) for SQ-LNS as incremental cost divided by DALYs averted. We performed all analyses at the draw-specific level and presented results summarized across all draws as mean and 95 percent uncertainty intervals (UIs) unless otherwise noted. ICERs are reported as summaries of draw-specific cost to impact ratios rather than the ratio of summarized costs and impacts.

All costs and impacts are represented as annual values among the population six months to five years of age after the effects of the SQ-LNS program have reached full saturation throughout the under-five year population (i.e. operating at scale for at least 4.5 years) to represent fully scaled annual impacts and costs. We did not perform discounting of costs or impact of our intervention due to a time horizon of only one year in our model. We did not follow a pre-specified analysis protocol or consult with external stakeholders, patients, or public groups in the development of this analysis. All analyses were conducted in python.

### Analysis of Threshold-Based Targeting Algorithms

We modeled threshold-based targeting algorithms by summing the incremental costs and DALYs averted in all Admin1 locations in a given country that met the threshold criteria and calculated the resulting ICER. We considered threshold-based targeting approaches based on high GHI, wasting, stunting, and wasting, stunting, and anemia burdens among the location-specific under-five population. We defined high wasting as prevalence > 10%, high stunting as prevalence > 20%, and high anemia as prevalence > 40% as suggested by the SQ-LNS Operational Guidance [8]. We additionally considered thresholds based on GHI score corresponding with “serious” (> 20) and “alarming” (> 35) [20].

### Analysis of Priority-Based Targeting Algorithms

To model priority-based targeting algorithms, we ordered Admin1 locations in ascending order of a given targeting indicator within each country. We calculated the total population coverage, incremental costs, DALYs averted, and ICERs of iteratively including an additional Admin1 location from the priority-ordered list. To enable direct comparison between priority-based targeting indicators, we performed linear interpolation to obtain values specific to 30 and 60 percent national population coverage and to the incremental cost of threshold-based targeting approaches to enable comparison between threshold- and priority-based algorithms.

This process was performed for all targeting indicators as well as for mean and draw-specific values of subnational SQ-LNS ICERs. The draw-specific SQ-LNS ICER indicator was used as the maximum potential targeting efficiency benchmark in our analysis. We also considered a benchmark of an untargeted national scale-up strategy in which impacts scaled directly with costs and coverage and ICERs were constant across coverage levels. For each indicator as well as mean model-predicted cost-effectiveness, we assessed cost-effectiveness as percent improvement relative to the untargeted benchmark (equation 1) as well as cost-effectiveness as a percentage between the maximum targeting efficiency (100%) and untargeted (0%) benchmarks (Equation 2) at a given coverage level.

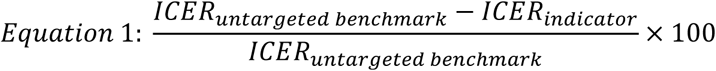

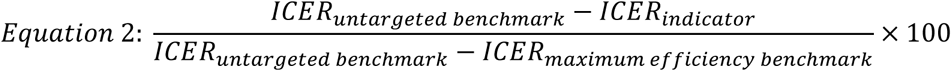

We performed a sensitivity analysis in which we considered alternative age groups and severity thresholds for our targeting indicator values. Alternative age groups included six to 24 months for mortality risk and wasting, 24-60 months for stunting, and under-five for all individual measures included in the composite ranking indicator. Alternative severity thresholds were severe for child growth failure measures (z-scores < −3) and anemia (hemoglobin <90 g/L among neonates and < 70 g/L otherwise), and total (z-scores < −2) for wasting and stunting measures to inform the composite ranking indicator. We assessed the performance of these alternative targeting indicator definitions relative to the default definition in the same manner as we compared performance of targeting indicators to the maximum impact potential benchmark.

## Results

### Subnational-level results

Across Admin1 locations in Ethiopia, mortality risk ranged from 16 deaths per 1,000 live births in Addis Ababa to 69 in Somali, wasting prevalence from 5.2% in Addis Ababa to 20.2% in Somali, and GHI scores from 11.4 in Addis Ababa to 37.9 in Afar. Values for all targeting indicators for Admin1 locations in Ethiopia are shown in Table 1 as an illustrative example. Analogous values for Nigeria and Pakistan are provided in Appendix 1. Baseline values from our simulation had excellent agreement with verification targets (Lin’s Concordance Correlation Coefficient > 0.9, Appendix 3).

**Table 1.**
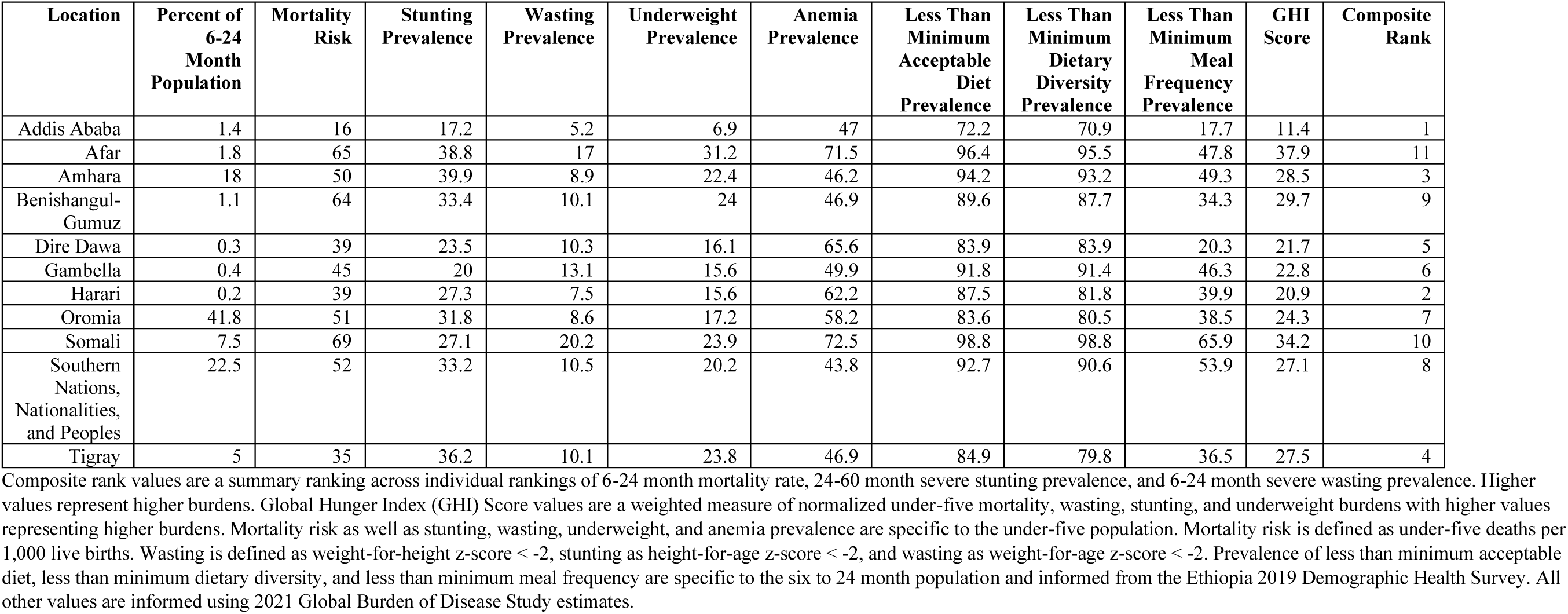
Targeting Indicator Values in Ethiopian First-Level Administrative Divisions.

At the subnational level, SQ-LNS ICERs ranged from $226 (UI: 143 to 426) in Somali to $1,580 (UI: 896 to 2,590) in Addis Ababa for Ethiopia, $77.7 (UI: 44.3 to 161) in Sokoto to $1,260 (UI: 499 to 3,160) in Osun for Nigeria, and $308 (UI: 202, 476) in Balochistan to $2,470 (UI: 1,650 to 3,620) in Islamabad Capital Territory in Pakistan. Scaling up SQ-LNS in all Admin1 locations would result in 217 thousand (UI: 110 to 313) DALYs averted and $90.9 million (UI: 84.7 to 95.9) incremental costs, with an ICER of $458 (UI: 291 to 863) per DALY averted in Ethiopia; 1.65 million (UI: 0.569 to 2.37) DALYs averted and $288 million (UI: 279 to 298) incremental costs, with an ICER of $227 (UI: 124 to 513) in Nigeria; and 473 thousand (UI: 278 to 647) DALYs averted and $240 million (UI: 230 to 251) incremental costs, with an ICER of $544 (UI: 366 to 878) in Pakistan. Table 2 shows Admin1 location-specific simulated estimates of baseline DALYs among the six to 59 month population as well as ICERs, DALYs averted, and incremental costs of SQ-LNS for all Admin1 locations in Ethiopia (analogous values for Nigeria and Pakistan are provided in Appendix 1). Incremental costs spent on AM treatment are negative as SQ-LNS reduced wasting incidence and associated AM treatment needs.

**Table 2:** Subnational Small-Quantity Lipid-Based Nutrient Supplementation (SQ-LNS) Health Impacts, Costs, and Cost-Effectiveness in Ethiopia.

| Location | Annual DALYs at baseline among the 6-59 month population | Annual DALYs averted | Incremental cost-effectiveness ratio | Total incremental cost | Incremental cost: SQ-LNS | Incremental cost: acute malnutrition treatment |
| --- | --- | --- | --- | --- | --- | --- |
|  |  |  | (USD / DALY averted) | (Millions of USD / year) |  |  |
| Addis Ababa | 28,400 (22,800, 35,200) | 1,010 (564, 1,650) | 1,580 (896, 2,590) | 1.42 (1.32, 1.48) | 1.46 (1.37, 1.52) | -0.0460<br>(-0.0678, -0.0243) |
| Afar | 156,000 (129,000, 195,000) | 6,790 (4,100, 10,800) | 248 (143, 376) | 1.56 (1.47, 1.67) | 1.71 (1.60, 1.78) | -0.150<br>(-0.225, -0.0594) |
| Amhara | 749,000 (636,000, 893,000) | 30,600 (16,400, 46,300) | 614 (374, 1,060) | 16.6 (15.6, 17.8) | 17.4 (16.2, 18.1) | -0.726<br>(-1.25, -0.135) |
| Benishangul-Gumuz | 86,900 (74,700, 106,000) | 3,070 (1,070, 4,530) | 391 (228, 942) | 1.02 (0.947, 1.08) | 1.08 (1.01, 1.12) | -0.0561<br>(-0.0833, -0.0285) |
| Dire Dawa | 16,400 (13,900, 18,900) | 544 (314, 796) | 579 (377, 906) | 0.296 (0.272, 0.316) | 0.318 (0.297, 0.331) | -0.0222<br>(-0.0359, -0.00711) |
| Gambella | 18,800 (15,000, 24,400) | 583 (234, 954) | 647 (323, 1,330) | 0.313 (0.289, 0.336) | 0.340 (0.318, 0.354) | -0.0269<br>(-0.0385, -0.0175) |
| Harari | 6,110 (4,730, 7,740) | 291 (147, 465) | 665 (377, 1,210) | 0.173 (0.162, 0.182) | 0.183 (0.171, 0.190) | -0.00948<br>(-0.0149, -0.00595) |
| Oromia | 2,130,000 (1,780,000, 2,520,000) | 83,000 (38,300, 128,000) | 524 (300, 1,100) | 38.3 (35.9, 40.2) | 40.3 (37.7, 41.8) | -1.97<br>(-2.84, -1.18) |
| Somali | 670,000 (567,000, 790,000) | 30,500 (15,100, 45,700) | 226 (143, 426) | 6.29 (5.72, 6.93) | 7.06 (6.62, 7.35) | -0.776<br>(-1.18, -0.413) |
| Southern Nations, Nationalities, and Peoples | 1,460,000 (1,220,000, 1,700,000) | 55,600 (26,200, 81,400) | 400 (251, 776) | 20.2 (18.6, 21.4) | 21.7 (20.3, 22.6) | -1.50<br>(-2.07, -1.06) |
| Tigray | 153,000 (130,000, 178,000) | 5,320 (2,740, 7,210) | 938 (662, 1,700) | 4.60 (4.30, 4.83) | 4.81 (4.50, 5.00) | -0.217<br>(-0.306, -0.153) |
| **National** | **5,480,000 (4,640,000, 6,440,000)** | **217,000 (110,000, 313,000)** | **458 (291, 863)** | **90.9 (84.7, 95.9)** | **96.4 (90.1, 100)** | **-5.50**<br>**(-7.58, -4.04)** |

DALY: Disability-adjusted life year; USD: 2021 United States Dollar; SQ-LNS: small-quantity lipid-based nutrient supplementation

All targeting indicators had negative correlation coefficients with mean SQ-LNS ICERs, at the subnational level in all modeled countries, indicating greater cost-effectiveness in areas with higher burden. Correlations were assessed using Spearman’s rank (two-tailed) and results were statistically significant at α = 0.05 without correction for multiple comparisons except anemia and less than minimum meal frequency prevalence in all modeled countries, mortality risk in Pakistan, and stunting prevalence in Ethiopia (results not shown). Scatterplots of each targeting indicator assessed with mean model-predicted SQ-LNS ICER are provided in Appendix 2.

### Threshold-based targeting results

Threshold-based targeting according to subnational locations with high wasting, stunting, and anemia burdens resulted in 38.6% population coverage with $34.3 (UI: 31.7 to 36.5) million annual incremental costs, 102,000 (UI: 52,100 to 143,000) annual DALYs averted, and an ICER of $363 (UI: 243 to 666) in Ethiopia. In Nigeria, population coverage was 39% with $109 million (UI: 106 to 113) annual incremental costs, 1,046,000 annual DALYs averted (UI: 363,000 to 1,488,000), and an ICER of $135 (UI: 74.2 to 305). In Pakistan, population coverage was 47.8% with $115 million annual incremental costs (UI: 109 to 119), 256,000 annual DALYs averted (UI: 161,000 to 348,000), and an ICER of $474 (UI: 329 to 718). Results specific to additional thresholds are provided in Appendix 1. This strategy was 20% (UI: 12 to 27) more cost-effective than the untargeted national scale-up strategy benchmark in Ethiopia and 40% (UI: 38 to 43) more cost-effective in Nigeria. Cost-effectiveness of this targeted strategy did not significantly differ from the untargeted benchmark at the 95% uncertainty level in Pakistan (12% more cost-effective, UI: −2.9 to 25).

### Priority-based targeting algorithm results

Figure 1 displays the incremental cost, impact, and cost-effectiveness by program coverage in each country for the maximum targeting impact potential benchmark. Marked points in this figure indicate the point at which full coverage of a given subnational location is achieved; blue lines represent prioritization at the draw-level and red lines represent prioritization based on the average across all modeled draws and display mean impacts and ICERs of that prioritization.

**Figure 1:**
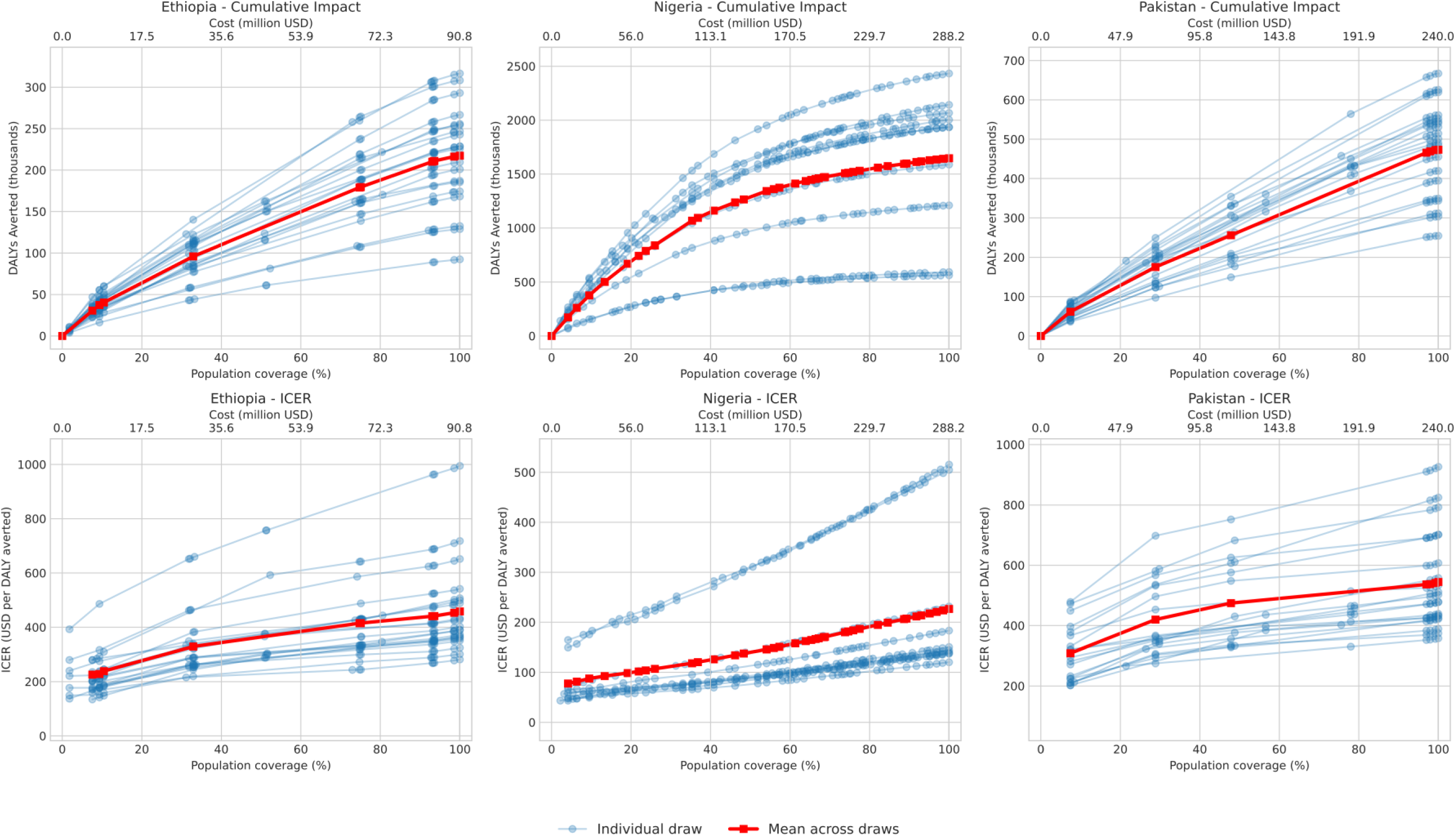
Impacts, Costs, and Cost-Effectiveness by Population Coverage with Priority-Based Targeting Using Model-Predicted Cost-Effectiveness as a Targeting Indicator <u>Footnote</u>: DALY: disability-adjusted life year; ICER: incremental cost-effectiveness ratio; USD: 2021 United States Dollars. Marked points in this figure indicate the point at which full coverage of a given subnational location is achieved; blue lines represent prioritization at the draw-level and red lines represent prioritization based on the average across all modeled draws.

As shown in Figure 2, a priority-based targeting algorithm using mean model-predicted cost-effectiveness resulted in greater DALYs averted relative to the untargeted uniformly distributed benchmark at the same level of spending. This finding was robust to modeled uncertainty and total population coverage of SQ-LNS in Nigeria and Ethiopia. In Pakistan, the mean impact of this targeted scale-up strategy was greater than an untargeted strategy across all coverage levels, but only differed from the untargeted strategy at the 95% uncertainty level with coverage values less than 40% and greater than 85%.

**Figure 2:**
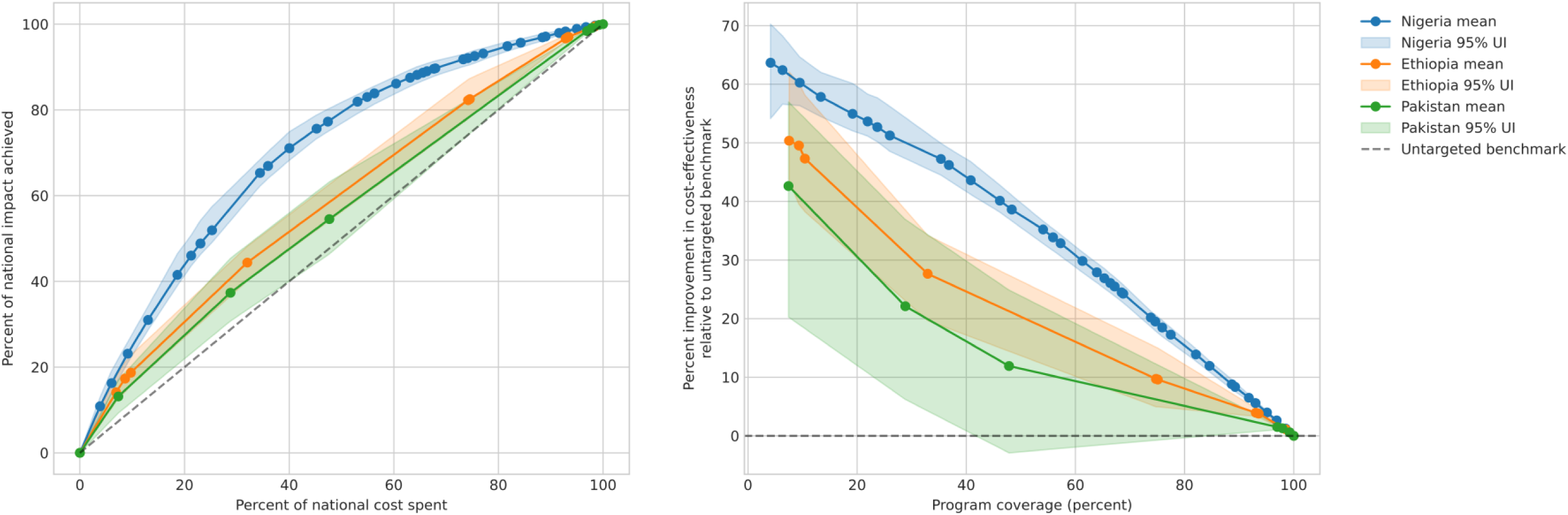
Performance of Priority-Based Targeting Using Mean Model-Predicted Cost-Effectiveness <u>Footnote:</u> UI: Uncertainty interval. Impact measured in disability-adjusted life years (DALYs) averted by small-quantity lipid-based nutrient supplementation (SQ-LNS). Total cost represents incremental cost of SQ-LNS in 2021 USD. Percent of total cost spent and percent of total impact achieved represent a percent of costs and impacts associated with scaling-up SQ-LNS in all Admin1 subnational locations within a given country. Cost-effectiveness measured as the incremental cost-effectiveness ratio (ICER) of SQ-LNS in 2021 United States Dollars (USD) per DALY averted. Population coverage represents proportion of the 6-24 month population residing in an Admin1 location with scaled-up SQ-LNS. Marked points in this figure indicate the point at which full coverage of a given subnational location is achieved.

In Ethiopia, priority-based targeting using mean model-predicted cost-effectiveness, composite rank, mortality risk, less than minimum dietary diversity prevalence, and less than minimum meal frequency prevalence were more cost-effective than the untargeted benchmark at 30% and 60% population coverage at the 95% uncertainty level (Figure 3). In Nigeria, priority-based targeting with all indicators was better than the untargeted benchmark at the 95% uncertainty level at 30% and 60% coverage except minimum meal frequency prevalence. Priority-based targeting with all indicators was more cost-effective than the untargeted benchmark at 30% population coverage in Pakistan and none were significantly more cost-effective than the untargeted benchmark at 60% population coverage except less than minimum acceptable meal frequency prevalence.

**Figure 3:**
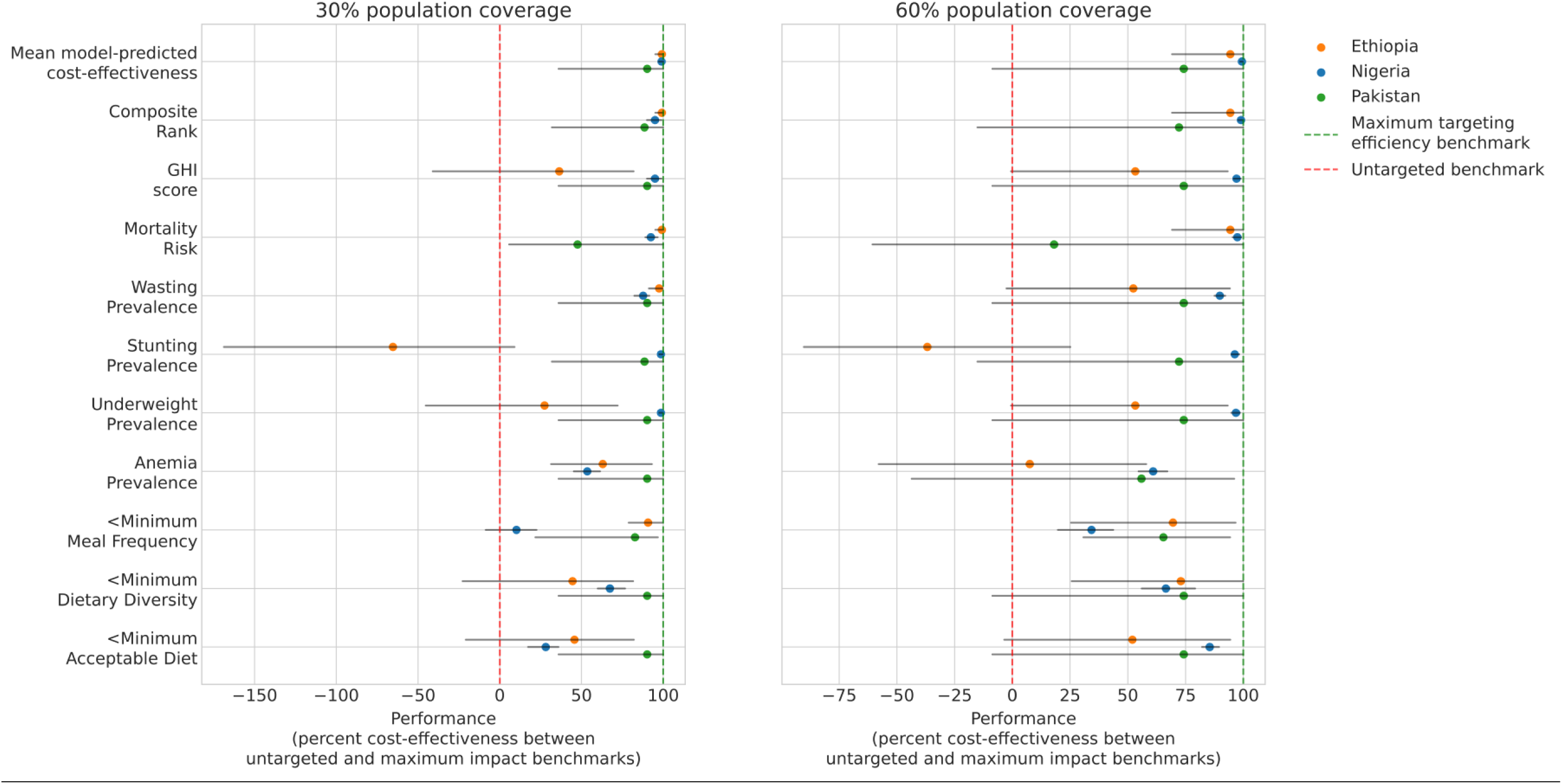
Performance of Priority-Based Targeting Algorithms by Targeting Indicator at 30 and 60 Percent Population Coverage <u>Footnote:</u> GHI: Global Hunger Index; Composite rank is the overall ranking across Admin1 locations within a given country across mortality, wasting, and stunting burdens. Error bars represent 95% uncertainty intervals. The maximum targeting efficiency benchmark represents the cost-effectiveness of targeting SQ-LNS to the given percent of the population with the greatest subnational model-predicted cost-effectiveness at the draw-level. The untargeted benchmark represents a uniformly distributed national scale-up of SQ-LNS to the given population coverage level. “Performance” values are calculated as difference in small-quantity lipid-based nutrient supplementation (SQ-LNS) incremental cost-effectiveness ratios (ICERs) with the untargeted benchmark and the given targeting indicator divided by the difference in ICERs with the evenly distributed and maximum impact benchmarks multiplied by 100 at the draw level and are shown as summaries across modeled draws.

The mean model-predicted cost-effectiveness indicator resulted in the greatest cost-effectiveness of all priority-based indicators, closely followed by the composite ranking indicator in all modeled locations at both 30% and 60% population coverage. Relative the untargeted benchmark, the mean model-predicted cost-effectiveness was estimated to achieve 49.3% (UI: 47.0 to 53.1) and 30.8% (UI: 28.8 to 32.3) greater cost-effectiveness in Nigeria at 30% and 60% population coverage respectively, 28.8% (UI: 21.3 to 35.3) and 13.9% (UI: 9.3 to 18.8) in Ethiopia, and 21.2% (UI: 5.4 to 35.9) and 8.2% (UI: −1.0 to 16.9) in Pakistan. The composite ranking indicator resulted in cost-effectiveness 47.3% (UI: 44.4 to 50.1) and 30.7% (UI: 28.6 to 32.1) greater than the untargeted benchmark at 30% and 60% population coverage in Nigeria, 28.8% (UI: 2.3 to 35.3) and 13.9% (UI: 9.3 to 18.8) in Ethiopia, and 21.0% (UI: 5.0 to 36.0) and 8.0% (UI: −1.6 to 17.2) in Pakistan. While other targeting indicators closely follow the performance of model-predicted cost-effectiveness in individual countries, no other indicators do so in all three modeled locations at both 30 and 60 percent coverage (Figure 3). Figures representing priority-based targeting indicator performance across all population coverage levels in each country are provided in Appendix 4. Targeting indicators that performed significantly worse than the untargeted benchmark at the 95% uncertainty level in any modeled location at any coverage level included stunting prevalence in Ethiopia and less than minimum acceptable diet and less than minimum meal frequency prevalence in Nigeria.

Compared to threshold-based targeting using high wasting, stunting, and anemia, priority-based targeting with mean model-predicted cost-effectiveness and composite ranking indicators offered greater impacts for the same cost with 8.3% (UI: 5.0 to 15) and 7.0% (UI: 4.4 to 12) more DALYs averted in Nigeria, and 4.6% (UI: 1.4 to 7.3) more in Ethiopia for both indicators. There was no difference between these approaches in Pakistan (0%, UI: 0 to 0).

### Sensitivity analyses

#### Age group of targeting indicator population

Averaged across modeled locations, there were no significant differences in the performance of targeting indicators using alternative age groups or severity thresholds when averaged over coverage levels less than 50%. Results were suggestive that severe stunting prevalence among the 24-60 month population, severe anemia among the under-five population, and severe underweight among the under-five population performed better than the default indicators, but these findings were not certain at the 95% level, with 6.4% (UI: −7.9 to 20), 7.7% (UI: −0.28 to 19), and 6.1% (UI: −1.4 to 19) greater cost-effectiveness respectively. In Ethiopia, severe underweight prevalence performed significantly better than underweight prevalence among the under-five population with 8.9 (UI: 0.0 to 27) greater cost-effectiveness.

#### SQ-LNS Effect Modification

SQ-LNS effect modification by wasting burden resulted in decreased ICERs in subnational locations with high wasting burdens and increased ICERs in locations with low burdens, resulting in larger impacts of subnational targeting across all targeting strategies. Relative the untargeted benchmark, the mean model-predicted cost-effectiveness indicator was estimated to achieve 57.6% (UI: 54.3 to 59.6) and 35.1% (UI: 33.6 to 36.5) greater cost-effectiveness in Nigeria, 48.7% (UI: 37.9 to 63.4) and 31.9% (UI: 21.8 to 46.1) in Ethiopia, and 38.9% (UI: 27.3 to 50.5) and 22.0 (UI: 12.9 to 29.2) in Pakistan at 30% and 60% population coverage respectively. Full results specific to this sensitivity analysis are provided in Appendix 5.

## Discussion

Our analysis suggests that subnational targeting at the Admin1 level can increase cost-effectiveness of SQ-LNS programs relative to untargeted scale-up strategies. For example, targeting SQ-LNS coverage to 50% of the population with the highest subnational composite ranking across wasting, stunting, and mortality in Nigeria can achieve 78% (UI: 75 to 80) of total potential national impact at just 49% of the cost, lowering national-level ICERs by 38% (UI: 36 to 40) from $227 (UI: 124 to 513) to $142 (UI: 80 to 318). Sensitivity analysis suggests that these impacts could be even greater if SQ-LNS effects are greater in targeted areas, such as in areas with high wasting burden as suggested by evidence in Dewey et al. (2022) [6].

According to our results, targeted SQ-LNS programs can offer greater cost-effectiveness than untargeted programs in our modeled countries with the use of several different targeting strategies. Priority-based targeting with composite ranking offered the greatest cost-effectiveness of all strategies assessed, with modest performance improvements relative to a threshold-based targeting approach using wasting, stunting, and anemia burdens. Threshold-based algorithms offer greater ease of use than priority-based algorithms that may justify this performance deficit for general guidelines. However, priority-based algorithms offer more flexibility in intended program coverage and may offer an alternative approach in specific cases when available program resources are not well matched to those required to achieve the coverage level implied by threshold-based approaches.

To our knowledge, this is the first analysis that evaluates the cost-effectiveness of subnational SQ-LNS targeting strategies. This analysis builds on the extensive work published on the SQ-LNS to date, integrating information from existing analyses of health impacts [2], costs [4,5,15], operational guidelines [8,10,21], and priority location identification [9]. Our analysis benefits from a detailed model of SQ-LNS intervention impact informed from robust GBD study estimates. Utilizing the GBD quantification of child growth failure’s effects on morbidity and mortality rather than directly applying the meta-analyzed mortality effects of SQ-LNS allows for modeled impacts that are more specific to the underlying epidemiology of a given subnational location.

There are several notable limitations to our model. First, it is an *in silico* model of predicted SQ-LNS impact that has not been externally validated and model-estimated cost-effectiveness could vary from true cost-effectiveness upon implementation. It is informed from 2021 estimates and does not consider the impact of SQ-LNS on any developmental outcomes nor any potential synergistic effects on coverage of services coupled with SQ-LNS distribution such as vaccinations or AM screenings despite evidence or suggestion of such effects [22–25]. While we benefit from the use of Monte Carlo uncertainty propagation in our health impact simulation, the number of draws utilized is limited and may not represent the full range of uncertainty expected. Additionally, our assumed intervention costs are informed from limited trial contexts and the cost of programmatic coverage may vary. We do not consider any uncertainty or subnational variation in intervention costs although costs related to personnel, distribution, and infrastructure are unlikely to be constant across subnational locations. Our model also does not consider feasibility of scaling SQ-LNS in any of our modeled locations nor feasibility of targeting strategy implementation.

Our analysis is additionally limited in that it did not consider targeting based on food insecurity as recommended by the WHO [10] and only assessed three countries and does not represent all locations that could benefit from SQ-LNS. Additionally, the division of subnational Admin1 locations assessed may not align with the geographical units policy makers may consider for SQ-LNS programming. For instance, Oromia makes up more than 40% of the six to 24 month population in Ethiopia and burden within this population is not likely to be homogenous. Targeting assessment at the second- or third-level administrative district may prove to have more utility, especially in countries with few Admin1 districts like Pakistan. Additionally, our analysis considered only geographic-level targeting and did not consider individual-based targeting strategies. While there are advantages to geographic-based approaches such as ease of implementation, individual-based targeting may enable increased impact per child covered by further refining the population who will benefit the most [26,27]. For example, Ruel et al. (2008) [28] conducted a study in which SQ-LNS was targeted to underweight children, although cost-effectiveness of this strategy was not assessed. There has been more recent consideration of targeting SQ-LNS to children upon recovery from severe AM to reduce relapse risk [29,30].

Our findings validate the SQ-LNS location prioritization approach using a custom ranking metric across mortality, wasting, and stunting published by Noori et al. (2025) [9] by linking this metric to location-specific cost-effectiveness of SQ-LNS in a detailed health impact model. Sensitivity analysis revealed similar performance of this indicator informed using total prevalence among the under-five population compared to the more specific values used by Noori et al. (2025) [9] of severe prevalence among narrower age groups. While the most relevant data should be prioritized when available, this suggests that using convenience data may be an acceptable alternative for policymakers considering this prioritization approach. Our composite rankings broadly agreed with those of Noori et al. (2025) [9] at the subnational level in Nigeria and Pakistan (not assessed for Ethiopia), sharing four of five top-priority locations in Nigeria and the top two in Pakistan, with more divergence further down the rankings.

SQ-LNS ICER estimates in our model have a wider range (mean estimates of $77.7 in Sokoto, Nigeria to $2,470 in Islamabad Capital Territory, Pakistan) than those found in the existing literature ($242 in Uganda [4] to $747 in Mali and $1,073 in Burkina Faso [5]). Reasons for ICERs higher than those observed in the literature are likely due to evaluation in populations that would not have been selected for SQ-LNS studies due to low predicted impact/cost-effectiveness. Reasons that our predicted ICERs are lower than those in the existing literature may include our model capturing the impact of SQ-LNS on mortality mediated through reductions in child stunting through five years of age rather than only during the period of SQ-LNS administration (6 to 24 months). Additionally, our model captures cost savings on AM treatment associated with averted cases of child wasting that partially offset the cost of SQ-LNS.

Future directions include integrating a targeted SQ-LNS intervention into the collection of interventions supported by the health impact and allocative efficiency model described in Bowman et al. (2025) [11] to assess its prioritization among alternative maternal and child nutrition interventions. Additionally, an exploration of individual-based SQ-LNS targeting algorithms would be a worthwhile extension of this work.

In conclusion, our analysis found targeting SQ-LNS at the Admin1 level can significantly improve cost-effectiveness in Nigeria and Ethiopia across all coverage levels and for low (<40%) and very high (>85%) coverage levels in Pakistan. Priority-based targeting with a composite ranking indicator across mortality, wasting, and stunting burdens had the greatest cost-effectiveness of all strategies assessed. Policy makers should consider targeting at the subnational level in the design of their SQ-LNS implementation strategies in addition to following the recommendations in existing guidelines [7,8,10,21].

## Data Availability

This analysis used publicly available estimates from the Global Burden of Disease (GBD) 2021 study, accessible via the Global Health Data Exchange (https://ghdx.healthdata.org/gbd-2021) under the IHME Free-of-Charge Non-Commercial User Agreement, and from the Nigeria, Ethiopia, and Pakistan Demographic and Health Surveys cited in the manuscript (references 17–19). SQ-LNS effect sizes on child wasting and stunting were obtained through personal correspondence with the authors of Dewey et al. (2021) [2]. The simulation inputs and code used to generate results for this analysis are available for the pregnancy model at https://doi.org/10.5281/zenodo.22262631 and for the child model at https://doi.org/10.5281/zenodo.22261085.

## Acknowledgements

We would like to acknowledge Osoti Osoti, MPH, Jomo Kenyatta University of Agriculture and Technology, for a contribution that led to the modeling decision to pause small-quantity lipid-based nutrient supplementation cost accrual during active acute malnutrition treatment.

## Author Contributions

AB and SL contributed to conceptualization and formal analysis. AB, SL, and AP contributed to data curation. AB led investigation. AB, SL, ZB, HJ, and JA contributed to validation. AB, SL, ZB, NB, and RM contributed to methodology. AB and ZB contributed to visualization. AB wrote the initial manuscript draft and all authors reviewed and edited. HJ, JA, and RM contributed to software. SL, RM, and AF contributed to project administration. AF led funding acquisition and supervision.

## Ethics approval

Not applicable. This modelling study used aggregate, publicly available population-level estimates and survey statistics (Global Burden of Disease 2021 estimates and Demographic and Health Survey reports); no primary data were collected and no individual-level or identifiable data were accessed.

## What is already known on this topic

Small-quantity lipid-based nutrient supplementation (SQ-LNS) has significant effects on child mortality, nutrition, and developmental outcomes. SQ-LNS is recommended for use in populations it is hypothesized to benefit, including those with high burdens of food insecurity, growth failure, nutritional deficiencies, and/or mortality.

## What this study adds

This study finds that targeting SQ-LNS to subnational populations with the greatest mortality, wasting, and stunting burdens could result in up to 50 percent more impact on child morbidity and mortality than an untargeted program of the same scale.

## How this study might affect research, practice or policy

This study strengthens the evidence to support existing recommendations to target SQ-LNS to specific populations and provides policymakers with additional evidence for translating these recommendations into practice.

## List of abbreviations

Admin1: First-level administrative division
AM: acute malnutrition
CGF: Child growth failure
DALY: Disability-adjusted life year
g/L: grams per liter
GBD: Global Burden of Disease
GHI: Global Hunger Index
HAZ: Height-for-age z-score
ICER: Incremental cost-effectiveness ratio
SQ-LNS: Small-quantity lipid-based nutrient supplementation
UI: uncertainty interval
WAZ: Weight-for-age z-score
WHO: World Health Organization
WHZ: Weight-for-height z-score
YLD: Years lived with disability
YLL: Years of life lost

