## Appendix for "Cost-effectiveness of subnational targeting of small-quantity lipid-based nutrient supplementation: A simulation model in Nigeria, Ethiopia, and Pakistan"

**List of appendices**


**A1:** Full location tables

**A2:** Targeting Indicator and SQ-LNS scatter plots

**A3:** Baseline validation of simulation relative to targeting data

**A4:** Priority-based targeting indicator performance across full coverage spectrum

**A5:** SQ-LNS effect modification results

### Appendix 1: Full location tables

Table A1-1: Nigeria Targeting Data

| Subnational | Percent of 6-24 Month Population | Mortality Risk | Stunting Prevalence | Wasting Prevalence | Underweight Prevalence | Anemia Prevalence | Less Than Minimum Acceptable Diet Prevalence | Less Than Minimum Dietary Diversity Prevalence | Less Than Minimum Meal Frequency Prevalence | GHI Score | Composite Rank |
|---|---|---|---|---|---|---|---|---|---|---|---|
| Abia | 1.8 | 70 | 17.2 | 9.1 | 16.2 | 64.0 | 92 | 87.4 | 44.6 | 22.6 | 21 |
| Adamawa | 1.5 | 119 | 32.5 | 8.8 | 20.6 | 59.6 | 97.9 | 94.3 | 73.5 | 32.6 | 25 |
| Akwa Ibom | 0.8 | 58 | 22.3 | 9.1 | 17.5 | 73.2 | 82.6 | 71.2 | 38.8 | 23.2 | 22 |
| Anambra | 2.5 | 53 | 13.1 | 7.0 | 10.4 | 68.6 | 81.5 | 72.3 | 24.7 | 16.4 | 7 |
| Bauchi | 3.9 | 142 | 48.6 | 9.7 | 32.8 | 68.2 | 99.1 | 97.3 | 64.1 | 44.2 | 30 |
| Bayelsa | 0.3 | 60 | 17.1 | 4.7 | 10.7 | 69.5 | 90.6 | 80.0 | 58.2 | 16.8 | 9 |
| Benue | 4.0 | 86 | 21.5 | 6.4 | 12.9 | 63.6 | 93.7 | 84.0 | 63.2 | 22.2 | 17 |
| Borno | 2.2 | 67 | 35.0 | 14.4 | 26.1 | 64.6 | 99.4 | 93.8 | 86.5 | 33.6 | 28 |
| Cross River | 0.8 | 54 | 18.6 | 6.7 | 13.1 | 73.0 | 79.1 | 48.5 | 57.4 | 18.8 | 10 |
| Delta | 1.6 | 70 | 16.9 | 7.3 | 13.2 | 69.9 | 88.6 | 78.7 | 31.7 | 20.2 | 12 |
| Ebonyi | 1.4 | 86 | 21.2 | 8.5 | 15.7 | 77.9 | 96.6 | 94.3 | 36.5 | 24.5 | 24 |
| Edo | 1.4 | 60 | 15.2 | 7.0 | 11.1 | 70.9 | 84.4 | 59.8 | 59.6 | 17.9 | 13 |
| Ekiti | 0.6 | 68 | 18.8 | 5.8 | 11.8 | 70.1 | 95.2 | 81.9 | 72.6 | 19.0 | 6 |
| Enugu | 1.6 | 44 | 11.1 | 4.5 | 6.8 | 65.0 | 84.3 | 79.4 | 37.5 | 12.2 | 1 |
| FCT (Abuja) | 2.5 | 64 | 18.8 | 5.7 | 11.9 | 67.2 | 88.7 | 84.7 | 49.4 | 18.7 | 2 |
| Gombe | 1.8 | 120 | 43.7 | 10.5 | 29.8 | 66.3 | 99.3 | 98.6 | 58.4 | 40.0 | 29 |
| Imo | 4.1 | 51 | 14.3 | 7.7 | 12.2 | 68.2 | 90.0 | 79.9 | 46.8 | 17.7 | 11 |
| Jigawa | 3.1 | 142 | 55.2 | 13.7 | 38.2 | 91.4 | 95.1 | 91.7 | 49.1 | 50.2 | 36 |
| Kaduna | 5.3 | 93 | 42.2 | 11.5 | 27.5 | 59.6 | 94.2 | 91.9 | 43.0 | 36.7 | 27 |
| Kano | 9.3 | 137 | 45.7 | 12.8 | 31.8 | 70.4 | 94.1 | 83.4 | 66.1 | 44.3 | 31 |
| Katsina | 5.8 | 113 | 51.9 | 13.0 | 35.0 | 67.8 | 95.1 | 87.4 | 68.4 | 44.9 | 34 |
| Kebbi | 2.7 | 124 | 48.8 | 13.7 | 37.4 | 78.2 | 99.6 | 98.3 | 74.8 | 46.6 | 33 |
| Kogi | 1.1 | 53 | 22.3 | 8.5 | 15.4 | 64.1 | 91.0 | 87.1 | 52.5 | 21.5 | 20 |
| Kwara | 1.2 | 47 | 27.0 | 8.5 | 17.6 | 69.1 | 100.0 | 96.0 | 66.2 | 22.9 | 16 |
| Lagos | 1.8 | 54 | 10.7 | 9.6 | 12.7 | 57.1 | 86.0 | 69.2 | 55.1 | 18.3 | 5 |
| Nasarawa | 2.6 | 73 | 28.8 | 6.6 | 18.1 | 71.8 | 85.4 | 72.7 | 45.2 | 25.0 | 19 |
| Niger | 5.7 | 86 | 31.5 | 9.9 | 21.3 | 71.4 | 98.3 | 95.4 | 79.1 | 30.1 | 26 |
| Ogun | 4.6 | 61 | 23.2 | 7.7 | 18.5 | 63.1 | 84.2 | 73.2 | 51.8 | 23.3 | 14 |
| Ondo | 1.3 | 66 | 20.2 | 6.4 | 13.8 | 71.9 | 99.2 | 84.2 | 86.6 | 20.4 | 15 |
| Osun | 1.5 | 45 | 20.2 | 7.8 | 14.8 | 65.7 | 87.6 | 70.6 | 60.1 | 19.5 | 8 |
| Oyo | 2.1 | 43 | 22.9 | 6.7 | 15.5 | 63.5 | 87.1 | 76.3 | 60.6 | 19.8 | 3 |
| Plateau | 5.1 | 68 | 32.6 | 5.8 | 17.9 | 64.0 | 97.2 | 91.0 | 75.2 | 24.9 | 18 |
| Rivers | 1.3 | 70 | 13.2 | 6.3 | 9.1 | 73.5 | 91.4 | 88.8 | 41.1 | 17.1 | 4 |
| Sokoto | 4.1 | 153 | 50.5 | 16.2 | 35.5 | 74.3 | 97.1 | 93.6 | 55.1 | 50.4 | 37 |
| Taraba | 4.0 | 115 | 37.5 | 7.3 | 19.5 | 66.7 | 94.5 | 91.4 | 59.0 | 32.0 | 23 |
| Yobe | 2.3 | 98 | 49.8 | 15.4 | 37.4 | 65.3 | 98.4 | 97.7 | 75.3 | 45.3 | 32 |
| Zamfara | 2.2 | 169 | 51.4 | 11.6 | 32.6 | 79.5 | 98.6 | 98.1 | 59.8 | 48.4 | 35 |

Table A1-1 footnote: Composite rank values are a summary ranking across individual rankings of 6-24 month mortality rate, 24-60 month severe stunting prevalence, and 6-24 month severe wasting prevalence. Higher values represent higher burdens. Global Hunger Index (GHI) Score values are a weighted measure of normalized under-five mortality, wasting, stunting, and underweight burdens with higher values representing higher burdens. Mortality risk as well as stunting, wasting, underweight, and anemia prevalence are specific to the under-five population. Mortality risk is defined as under-five deaths per 1,000 live births. Wasting is defined as weight-for-height z-score < -2, stunting as height-for-age z-score < -2, and wasting as weight-for-age z-score < -2. Prevalence of less than minimum acceptable diet, less than minimum dietary diversity, and less than minimum meal frequency are specific to the six to 24 month population and informed from the Ethiopia 2019 Demographic Health Survey. All other values are informed using 2021 Global Burden of Disease Study estimates.

Table A1-2: Nigeria subnational results

| Location | Disability adjusted life years at baseline among the 6-59 month population | Disability adjusted life years averted | Incremental cost-effectiveness ratio | Total incremental cost (Millions of USD) | Incremental cost: SQ-LNS | Incremental cost: acute malnutrition treatment |
|---|---|---|---|---|---|---|
| | | | (USD / DALY) | (Millions of USD) | | |
| Abia | 459,000 (399,000, 517,000) | 17,800 (6,570, 27,600) | 372 (196, 822) | 5.28 (5.16, 5.44) | 5.57 (5.36, 5.75) | -0.288 (-0.421, -0.175) |
| Adamawa | 630,000 (544,000, 692,000) | 26,100 (10,800, 43,700) | 211 (104, 425) | 4.44 (4.27, 4.61) | 4.59 (4.39, 4.77) | -0.145 (-0.223, -0.116) |
| Akwa Ibom | 145,000 (124,000, 172,000) | 6,180 (1,810, 10,600) | 528 (219, 1,310) | 2.29 (2.23, 2.38) | 2.40 (2.31, 2.48) | -0.112 (-0.161, -0.0841) |
| Anambra | 411,000 (353,000, 455,000) | 14,000 (3,180, 24,400) | 825 (312, 2,430) | 7.39 (7.13, 7.68) | 7.62 (7.32, 7.87) | -0.228 (-0.290, -0.126) |
| Bauchi | 2,360,000 (2,010,000, 2,690,000) | 126,000 (44,400, 171,000) | 111 (66.2, 268) | 11.2 (10.7, 11.6) | 11.6 (11.1, 12.0) | -0.431 (-0.588, -0.306) |
| Bayelsa | 58,600 (49,100, 67,800) | 1,970 (564, 3,070) | 601 (285, 1,610) | 0.862 (0.831, 0.889) | 0.882 (0.848, 0.906) | -0.0191 (-0.0341, -0.0143) |
| Benue | 952,000 (851,000, 1,030,000) | 38,200 (12,100, 64,400) | 423 (189, 1,020) | 11.9 (11.5, 12.4) | 12.3 (11.8, 12.7) | -0.346 (-0.475, -0.215) |
| Borno | 500,000 (422,000, 560,000) | 27,100 (8,450, 36,400) | 304 (169, 792) | 6.25 (6.03, 6.42) | 6.67 (6.42, 6.90) | -0.419 (-0.542, -0.300) |
| Cross River | 136,000 (125,000, 154,000) | 4,650 (1,930, 6,530) | 653 (374, 1,330) | 2.49 (2.40, 2.57) | 2.57 (2.47, 2.65) | -0.0714 (-0.107, -0.0529) |
| Delta | 314,000 (286,000, 338,000) | 10,000 (2,760, 17,600) | 681 (275, 1,730) | 4.69 (4.53, 4.86) | 4.84 (4.65, 5.01) | -0.153 (-0.218, -0.117) |
| Ebonyi | 322,000 (272,000, 359,000) | 13,500 (4,650, 21,400) | 385 (190, 884) | 4.02 (3.91, 4.12) | 4.19 (4.02, 4.31) | -0.172 (-0.252, -0.114) |
| Edo | 309,000 (263,000, 356,000) | 11,000 (2,930, 18,200) | 508 (219, 1,480) | 3.94 (3.79, 4.07) | 4.09 (3.93, 4.23) | -0.160 (-0.219, -0.114) |
| Ekiti | 120,000 (93,900, 139,000) | 3,580 (844, 6,540) | 792 (277, 2,170) | 1.79 (1.73, 1.83) | 1.84 (1.77, 1.89) | -0.0501 (-0.0636, -0.0341) |
| Enugu | 223,000 (198,000, 239,000) | 5,960 (2,190, 10,800) | 1,070 (479, 2,380) | 5.00 (4.82, 5.14) | 5.11 (4.91, 5.26) | -0.111 (-0.161, -0.0851) |
| FCT (Abuja) | 430,000 (348,000, 500,000) | 13,500 (3,880, 20,900) | 753 (346, 1,990) | 7.38 (7.09, 7.63) | 7.51 (7.20, 7.77) | -0.135 (-0.186, -0.0925) |
| Gombe | 830,000 (692,000, 955,000) | 45,600 (19,500, 69,500) | 138 (74.9, 269) | 5.11 (4.91, 5.33) | 5.34 (5.13, 5.50) | -0.228 (-0.345, -0.127) |
| Imo | 633,000 (546,000, 729,000) | 20,600 (7,030, 33,800) | 768 (361, 1,720) | 11.9 (11.5, 12.3) | 12.4 (11.9, 12.8) | -0.470 (-0.650, -0.307) |
| Jigawa | 1,910,000 (1,600,000, 2,160,000) | 115,000 (38,100, 174,000) | 106 (50.3, 236) | 8.94 (8.65, 9.23) | 9.45 (9.10, 9.73) | -0.505 (-0.685, -0.382) |
| Kaduna | 1,570,000 (1,380,000, 1,730,000) | 76,300 (23,400, 124,000) | 270 (121, 657) | 15.0 (14.6, 15.6) | 15.7 (15.1, 16.3) | -0.715 (-0.908, -0.547) |
| Kano | 4,870,000 (4,070,000, 5,490,000) | 228,000 (59,000, 347,000) | 173 (76.2, 465) | 26.5 (25.5, 27.6) | 27.8 (26.7, 28.9) | -1.37 (-1.91, -1.18) |
| Katsina | 2,890,000 (2,530,000, 3,210,000) | 169,000 (65,600, 243,000) | 118 (67.3, 254) | 16.2 (15.7, 16.9) | 17.1 (16.4, 17.7) | -0.919 (-1.34, -0.549) |
| Kebbi | 1,320,000 (1,110,000, 1,440,000) | 73,700 (24,700, 104,000) | 134 (72.9, 315) | 7.60 (7.36, 7.87) | 8.11 (7.81, 8.33) | -0.506 (-0.728, -0.414) |
| Kogi | 196,000 (180,000, 213,000) | 8,300 (2,430, 12,800) | 526 (250, 1,380) | 3.13 (3.04, 3.24) | 3.33 (3.21, 3.44) | -0.196 (-0.313, -0.144) |
| Kwara | 172,000 (144,000, 194,000) | 6,300 (1,500, 10,100) | 842 (358, 2,550) | 3.56 (3.45, 3.68) | 3.68 (3.54, 3.80) | -0.129 (-0.180, -0.0953) |
| Lagos | 236,000 (199,000, 254,000) | 6,960 (1,910, 11,500) | 1,040 (432, 2,820) | 5.09 (4.90, 5.30) | 5.35 (5.14, 5.53) | -0.258 (-0.366, -0.197) |
| Nasarawa | 555,000 (434,000, 628,000) | 22,900 (7,240, 35,300) | 467 (214, 1,110) | 7.68 (7.37, 7.92) | 7.90 (7.60, 8.13) | -0.225 (-0.319, -0.166) |
| Niger | 1,690,000 (1,470,000, 1,920,000) | 78,200 (19,300, 111,000) | 304 (146, 898) | 16.3 (15.7, 16.9) | 17.0 (16.4, 17.6) | -0.717 (-1.02, -0.504) |
| Ogun | 784,000 (675,000, 877,000) | 29,700 (6,670, 52,500) | 710 (261, 2,110) | 13.3 (12.8, 13.9) | 13.9 (13.3, 14.3) | -0.530 (-0.694, -0.433) |
| Ondo | 264,000 (230,000, 299,000) | 9,410 (2,850, 14,800) | 570 (260, 1,350) | 3.79 (3.67, 3.95) | 3.95 (3.79, 4.08) | -0.153 (-0.212, -0.0915) |
| Osun | 177,000 (157,000, 194,000) | 5,210 (1,410, 9,040) | 1,260 (499, 3,160) | 4.43 (4.26, 4.58) | 4.56 (4.38, 4.69) | -0.131 (-0.177, -0.0877) |
| Oyo | 309,000 (262,000, 348,000) | 10,600 (2,050, 17,400) | 993 (363, 3,330) | 6.23 (6.02, 6.45) | 6.39 (6.14, 6.59) | -0.163 (-0.250, -0.111) |
| Plateau | 936,000 (820,000, 1,040,000) | 35,700 (8,730, 48,400) | 635 (320, 1,840) | 15.3 (14.7, 15.8) | 15.7 (15.1, 16.3) | -0.386 (-0.495, -0.278) |
| Rivers | 285,000 (230,000, 326,000) | 7,980 (2,350, 13,400) | 674 (292, 1,790) | 3.83 (3.69, 3.95) | 3.91 (3.76, 4.03) | -0.0844 (-0.130, -0.0553) |
| Sokoto | 2,770,000 (2,450,000, 3,060,000) | 173,000 (70,200, 259,000) | 77.7 (44.3, 161) | 11.1 (10.7, 11.6) | 12.1 (11.6, 12.6) | -0.920 (-1.25, -0.685) |
| Taraba | 1,490,000 (1,190,000, 1,730,000) | 67,400 (27,200, 105,000) | 218 (113, 443) | 11.7 (11.4, 12.1) | 12.1 (11.6, 12.4) | -0.356 (-0.513, -0.224) |
| Yobe | 815,000 (689,000, 926,000) | 50,400 (18,300, 69,600) | 159 (91.4, 354) | 6.36 (6.21, 6.55) | 6.83 (6.58, 7.05) | -0.465 (-0.625, -0.322) |
| Zamfara | 1,640,000 (1,390,000, 1,810,000) | 88,000 (30,900, 137,000) | 91.8 (44.9, 208) | 6.31 (6.09, 6.52) | 6.65 (6.40, 6.84) | -0.340 (-0.445, -0.263) |
| **National** | 33,700,000 (30,600,000, 36,300,000) | 1,650,000 (569,000, 2,370,000) | 227 (124, 513) | 288 (279, 298) | 301 (289, 311) | -12.6 (-16.7, -9.94) |

DALY: Disability-adjusted life year; USD: 2021 United States Dollar; SQ-LNS: small-quantity lipid-based nutrient supplementation

Table A1-3: Pakistan targeting data

| Subnational | Percent of 6-24 Month Population | Mortality Risk | Stunting Prevalence | Wasting Prevalence | Underweight Prevalence | Anemia Prevalence | Less Than Minimum Acceptable Diet Prevalence | Less Than Minimum Dietary Diversity Prevalence | Less Than Minimum Meal Frequency Prevalence | GHI Score | Composite Rank |
|---|---|---|---|---|---|---|---|---|---|---|---|
| Azad Jammu & Kashmir | 1.3 | 49 | 19.5 | 6.7 | 14.3 | 55.7 | 83.2 | 72.8 | 38.3 | 19 | 2 |
| Balochistan | 7.4 | 62 | 51.8 | 22.1 | 38.4 | 59.3 | 92 | 85.6 | 38 | 46.5 | 7 |
| Gilgit-Baltistan | 1 | 46 | 37.6 | 5.7 | 19.1 | 56 | 72.5 | 63.9 | 30.3 | 24.4 | 5 |
| Islamabad Capital Territory | 0.8 | 15 | 22.2 | 9.7 | 9.8 | 55.3 | 76.7 | 62.5 | 27.8 | 16.2 | 1 |
| Khyber Pakhtunkhwa | 19 | 45 | 35.4 | 12.1 | 23.4 | 58 | 87.4 | 82.9 | 33.1 | 29.2 | 4 |
| Punjab | 49.1 | 62 | 29 | 9.7 | 23.3 | 51.2 | 85.4 | 75 | 37.7 | 27.9 | 3 |
| Sindh | 21.4 | 56 | 47.6 | 14.2 | 40 | 64 | 90.6 | 84.2 | 39.2 | 41.2 | 6 |

Table A1-2 footnote: Composite rank values are a summary ranking across individual rankings of 6-24 month mortality rate, 24-60 month severe stunting prevalence, and 6-24 month severe wasting prevalence. Higher values represent higher burdens. Global Hunger Index (GHI) Score values are a weighted measure of normalized under-five mortality, wasting, stunting, and underweight burdens with higher values representing higher burdens. Mortality risk as well as stunting, wasting, underweight, and anemia prevalence are specific to the under-five population. Mortality risk is defined as under-five deaths per 1,000 live births. Wasting is defined as weight-for-height z-score < -2, stunting as height-for-age z-score < -2, and wasting as weight-for-age z-score < -2. Prevalence of less than minimum acceptable diet, less than minimum dietary diversity, and less than minimum meal frequency are specific to the six to 24 month population and informed from the Ethiopia 2019 Demographic Health Survey. All other values are informed using 2021 Global Burden of Disease Study estimates.

Table A1-4: Pakistan subnational results

| Location | Disability adjusted life years at baseline among the 6-59 month population | Disability adjusted life years averted | Incremental cost-effectiveness ratio | Total incremental cost (Millions of USD) | Incremental cost: SQ-LNS | Incremental cost: acute malnutrition treatment |
|---|---|---|---|---|---|---|
| | | | (USD / DALY) | (Millions of USD) | | |
| Azad Jammu & Kashmir | 117,000 (87,200, 165,000) | 3,290 (1,880, 4,860) | 1,090 (657, 1,770) | 3.28 (3.14, 3.42) | 3.29 (3.15, 3.43) | -0.00712 (-0.00958, -0.00477) |
| Balochistan | 967,000 (769,000, 1,280,000) | 61,600 (37,200, 86,300) | 308 (202, 476) | 17.6 (16.8, 18.4) | 17.8 (17.0, 18.6) | -0.188 (-0.273, -0.110) |
| Gilgit-Baltistan | 78,100 (62,200, 106,000) | 3,460 (1,590, 5,410) | 737 (419, 1,480) | 2.26 (2.16, 2.34) | 2.26 (2.17, 2.36) | -0.00957 (-0.0155, -0.00405) |
| Islamabad Capital Territory | 17,800 (14,100, 21,400) | 797 (511, 1,140) | 2,470 (1,650, 3,620) | 1.86 (1.78, 1.93) | 1.87 (1.79, 1.94) | -0.00640 (-0.00864, -0.00345) |
| Khyber Pakhtunkhwa | 1,450,000 (1,160,000, 1,960,000) | 80,500 (51,600, 119,000) | 608 (377, 901) | 45.4 (43.4, 47.3) | 45.7 (43.8, 47.6) | -0.332 (-0.430, -0.205) |
| Punjab | 4,260,000 (3,490,000, 5,370,000) | 210,000 (102,000, 301,000) | 628 (390, 1,180) | 118 (113, 124) | 119 (114, 124) | -0.792 (-1.18, -0.389) |
| Sindh | 1,820,000 (1,390,000, 2,460,000) | 114,000 (71,500, 172,000) | 486 (295, 734) | 51.5 (49.2, 53.6) | 51.9 (49.7, 54.1) | -0.461 (-0.753, -0.262) |
| **National** | 8,720,000 (7,260,000, 11,200,000) | 473,000 (278,000, 647,000) | 544 (366, 878) | 240 (230, 251) | 242 (232, 252) | -1.80 (-2.41, -1.21) |

DALY: Disability-adjusted life year; USD: 2021 United States Dollar; SQ-LNS: small-quantity lipid-based nutrient supplementation

Table A1-5: Threshold-Based Targeting Algorithm Results

| Threshold | Ethiopia | | | | Nigeria | | | | Pakistan | | | |
|---|---|---|---|---|---|---|---|---|---|---|---|---|
| | **Coverage** | **Annual DALYs Averted (thousands)** | **Annual Incremental Cost (millions)** | **ICER** | **Coverage** | **Annual DALYs Averted (thousands)** | **Annual Incremen tal Cost (millions)** | **ICER** | **Coverage** | **Annual DALYs Averted (thousands)** | **Annual Incremental Cost (millions)** | **ICER** |
| High anemia (>40%) | 100 | 217 (110, 313) | 90.9 (84.7, 95.9) | 458 (291, 863) | 100 | 1,650 (569, 2,370) | 288 (279, 298) | 227 (124, 513) | 100 | 473 (278, 647) | 240 (230, 251) | 544 (366, 878) |
| High stunting (>20%) | 98.6 | 216 (109, 311) | 89.5 (83.4, 94.4) | 453 (288, 855) | 79.8 | 1,530 (531, 2,180) | 229 (222, 237) | 193 (106, 436) | 98.7 | 470 (276, 643) | 237 (226, 247) | 541 (363, 873) |
| High wasting (>10%) | 38.6 | 102 (52.1, 143) | 34.3 (31.7, 36.5) | 363 (243, 666) | 39 | 1,050 (363, 1,490) | 109 (106, 113) | 135 (74.2, 305) | 47.8 | 256 (161, 348) | 115 (109, 119) | 474 (329, 718) |
| High wasting + stunting + anemia | 38.6 | 102 (52.1, 143) | 34.3 (31.7, 36.5) | 363 (243, 666) | 39 | 1,050 (363, 1,490) | 109 (106, 113) | 135 (74.2, 305) | 47.8 | 256 (161, 348) | 115 (109, 119) | 474 (329, 718) |
| GHI > 20 (serious) | 98.6 | 216 (109, 311) | 89.5 (83.4, 94.4) | 453 (288, 855) | 79.6 | 1,540 (537, 2,200) | 228 (221, 236) | 191 (105, 430) | 97.9 | 469 (275, 642) | 235 (225, 245) | 538 (361, 868) |
| GHI > 35 (alarming) | 1.8 | 6.79 (4.10, 10.8) | 1.56 (1.47, 1.67) | 248 (143, 376) | 40.7 | 1,140 (410, 1,620) | 114 (111, 119) | 128 (71.4, 285) | 28.8 | 175 (109, 241) | 69.1 (66.1, 72.0) | 420 (283, 642) |

DALY: Disability-adjusted life year, ICER: Incremental cost-effectiveness ratio, GHI: Global Hunger Index Score, USD: 2021 United States Dollars

### Appendix 2: Targeting Indicator and SQ-LNS scatter plots

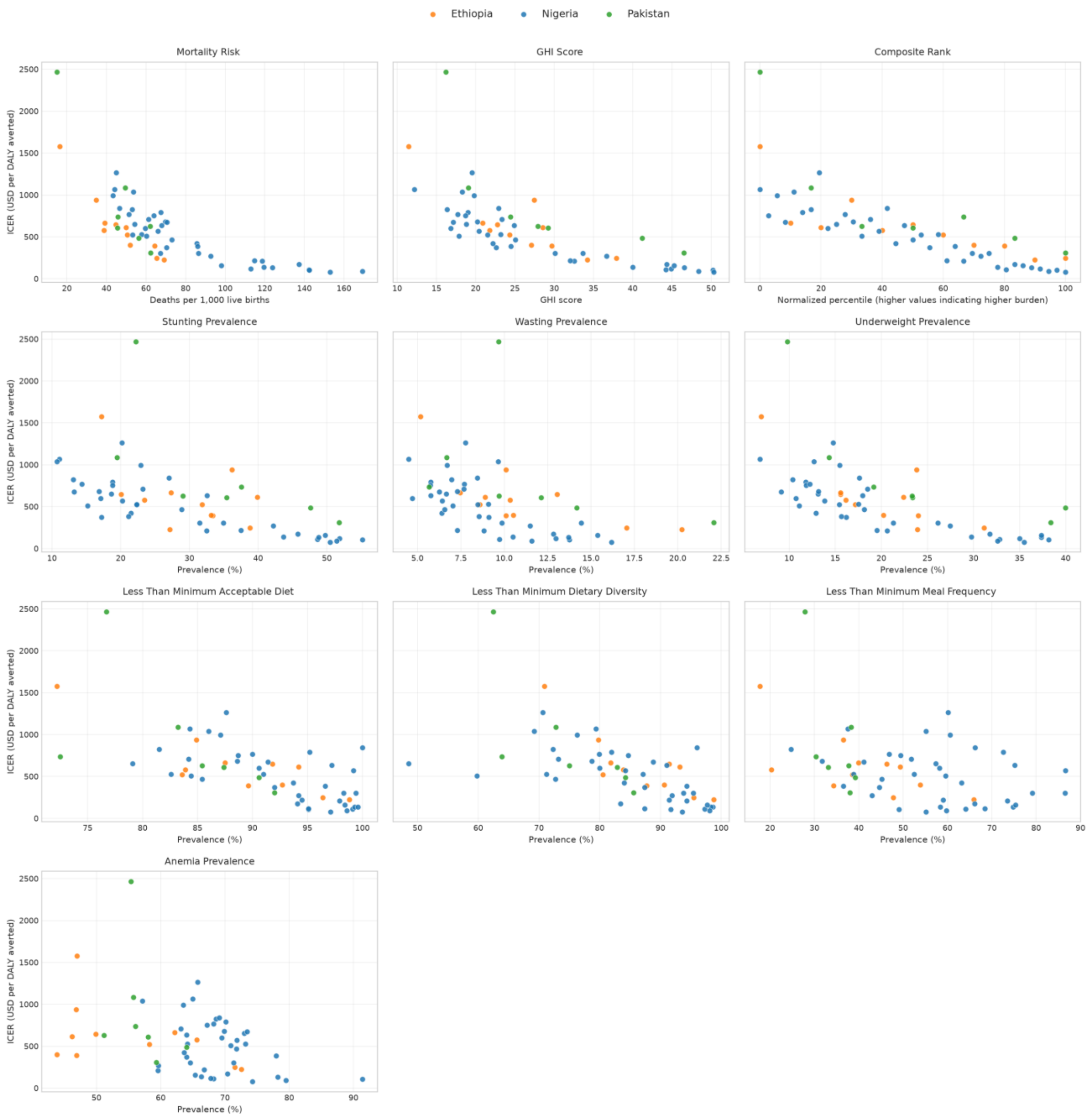


Composite rank values are a summary ranking across individual rankings of 6-24 month mortality rate, 24-60 month severe stunting prevalence, and 6-24 month severe wasting prevalence. Higher values represent higher burdens. Global Hunger Index (GHI) Score values are a weighted measure of normalized under-five mortality, wasting, stunting, and underweight burdens with higher values representing higher burdens. Mortality risk as well as stunting, wasting, underweight, and anemia prevalence are specific to the under-five population. Mortality risk is defined as under-five deaths per 1,000 live births. Wasting is defined as weight-for-height z-score < -2, stunting as height-for-age z-score < -2, and wasting as weight-for-age z-score < -2. Prevalence of less than minimum acceptable diet, less than minimum dietary diversity, and less than minimum meal frequency are specific to the six to 24 month population and informed from the Ethiopia 2019 Demographic Health Survey. All other values are informed using 2021 Global Burden of Disease Study estimates.

### Appendix 3: Baseline validation of simulation relative to targeting data

Mean simulated values for under-five mortality risk (under-five deaths per 1,000 live births), wasting prevalence, and stunting prevalence had substantial agreement with target indicator values at the subnational level (Lin's concordance correlation coefficients of 0.983 for mortality risk, 0.933 for wasting prevalence, and 0.996 for stunting prevalence).

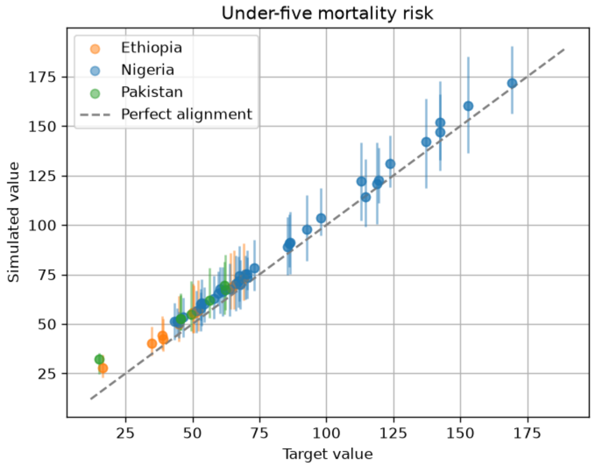


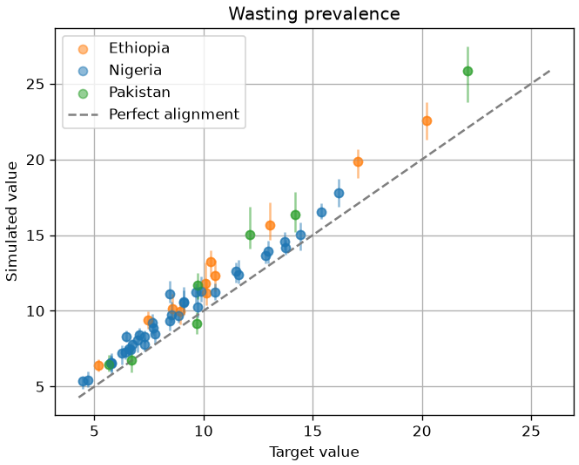


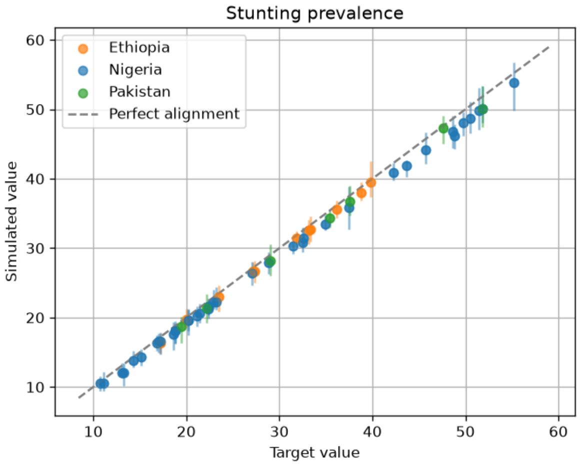

**Appendix 4: Priority-based targeting indicator performance across full coverage spectrum**

Figure A4-1: Priority-based targeting indicator cost-effectiveness relative to the untargeted benchmark

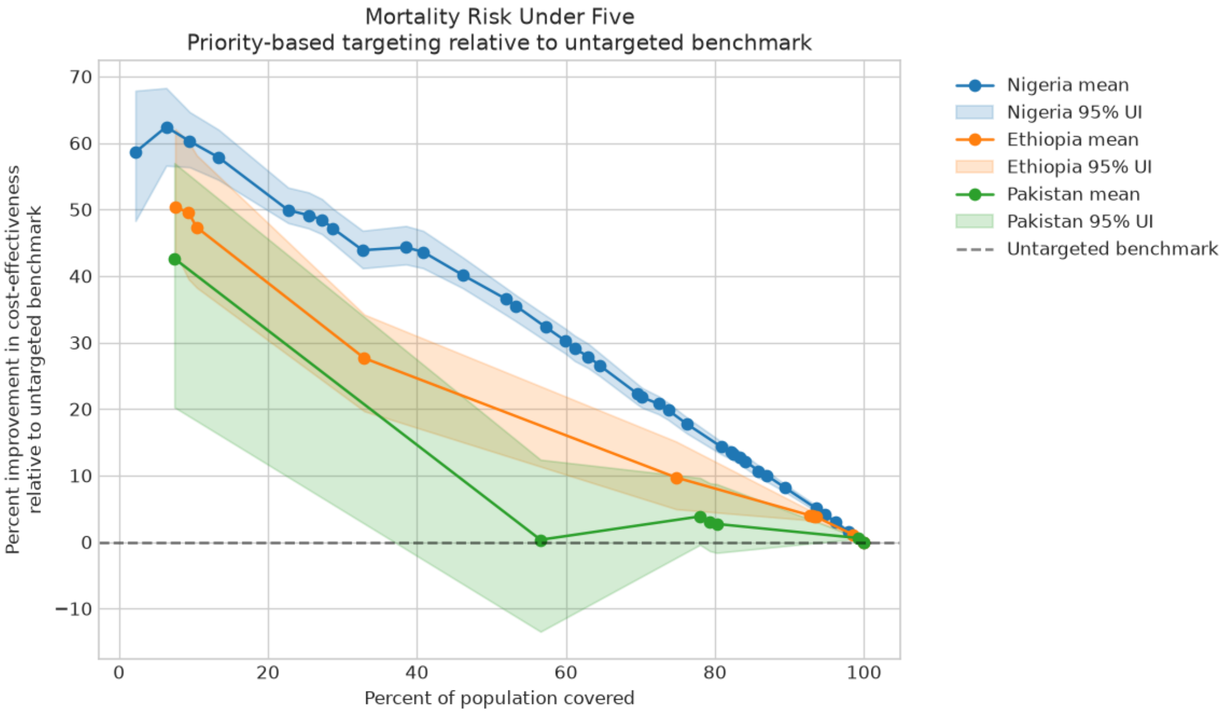


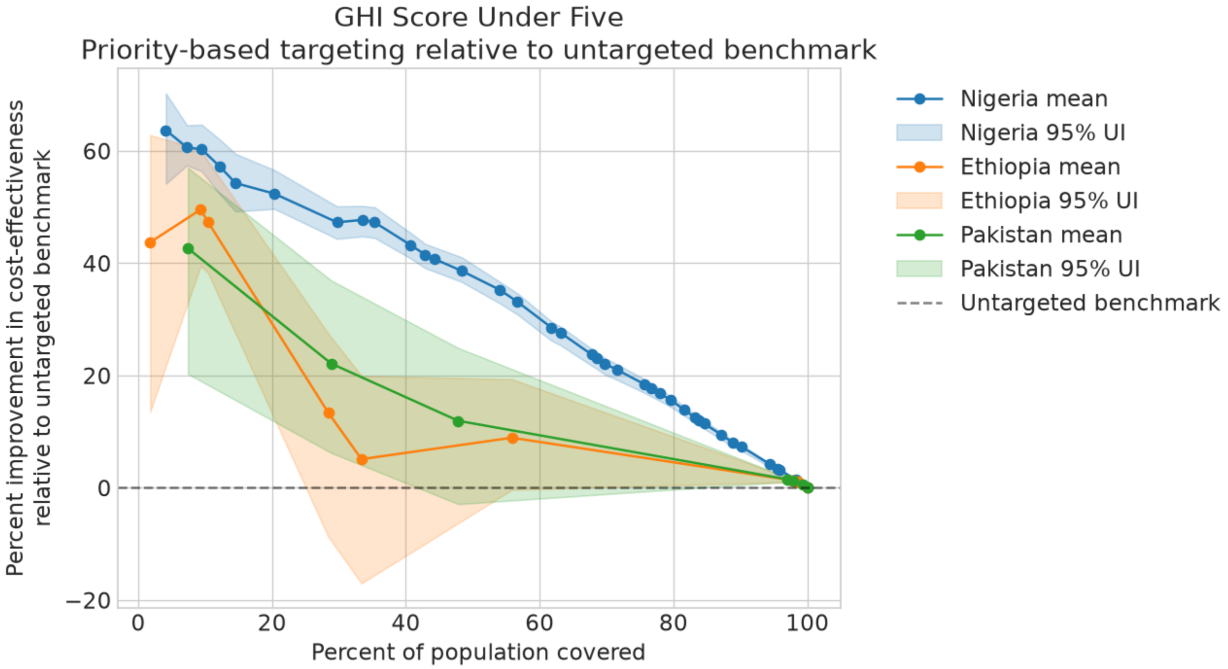

Figure A4-1: Priority-based targeting indicator cost-effectiveness relative to the untargeted benchmark (continued)

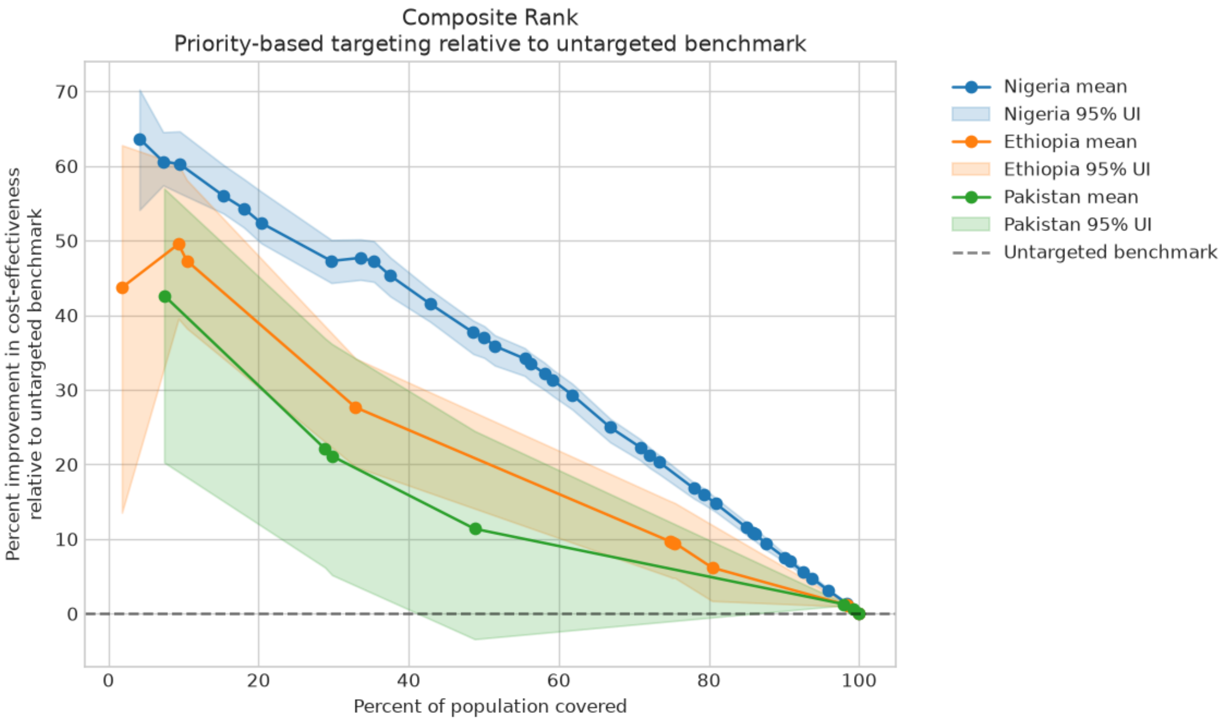


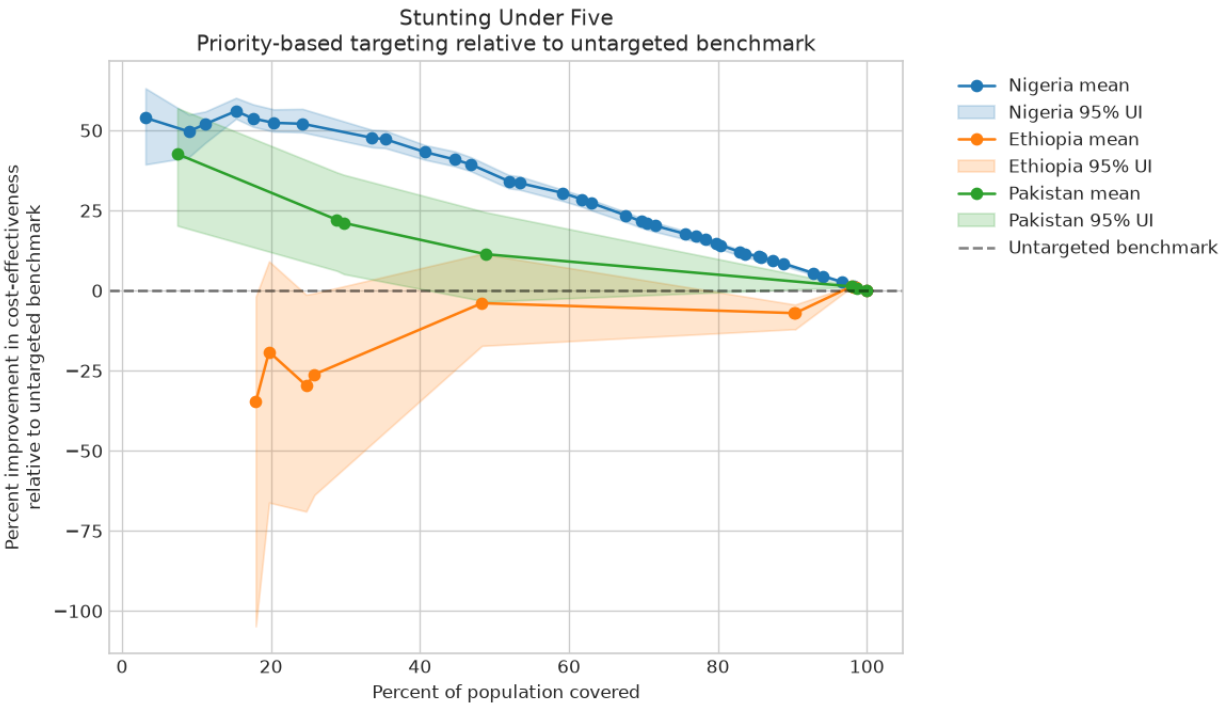

Figure A4-1: Priority-based targeting indicator cost-effectiveness relative to the untargeted benchmark (continued)

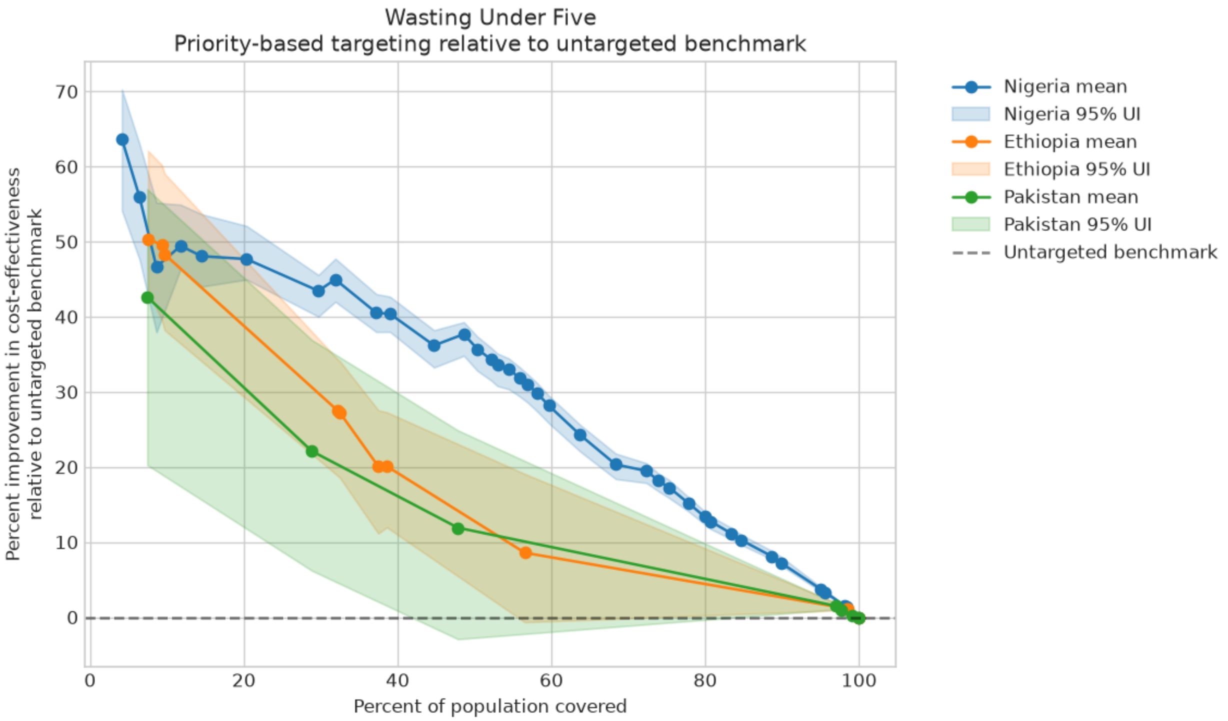


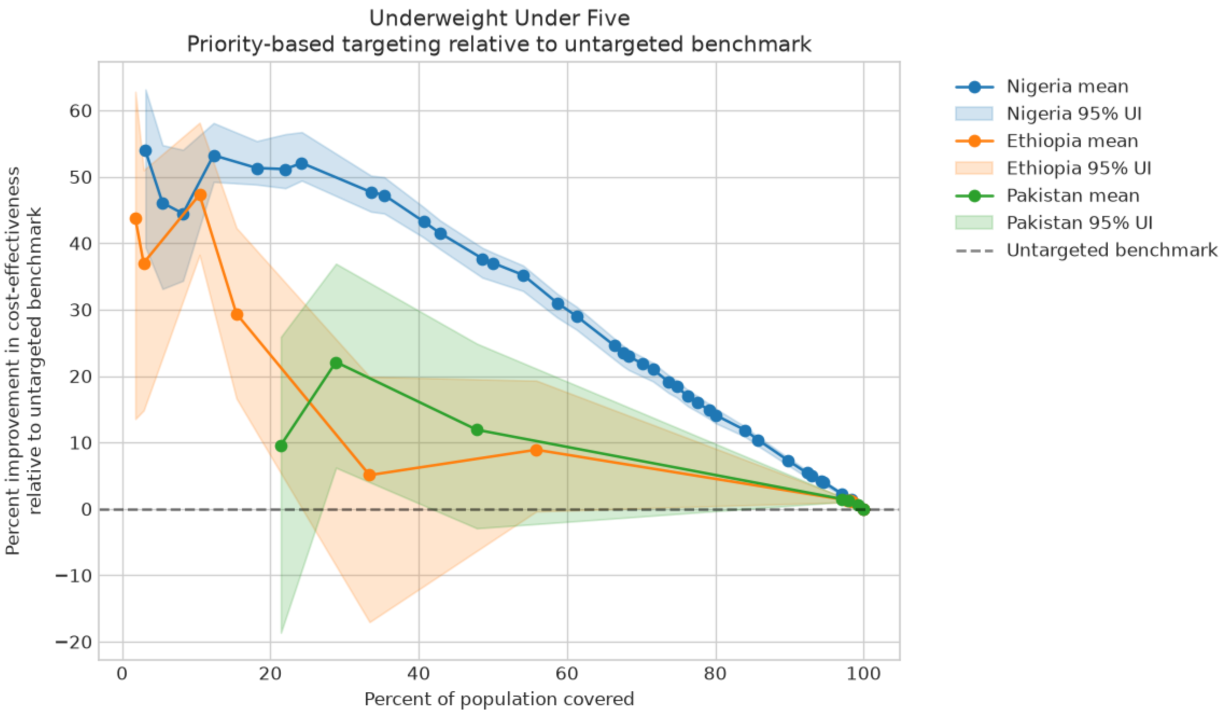

Figure A4-1: Priority-based targeting indicator cost-effectiveness relative to the untargeted benchmark (continued)

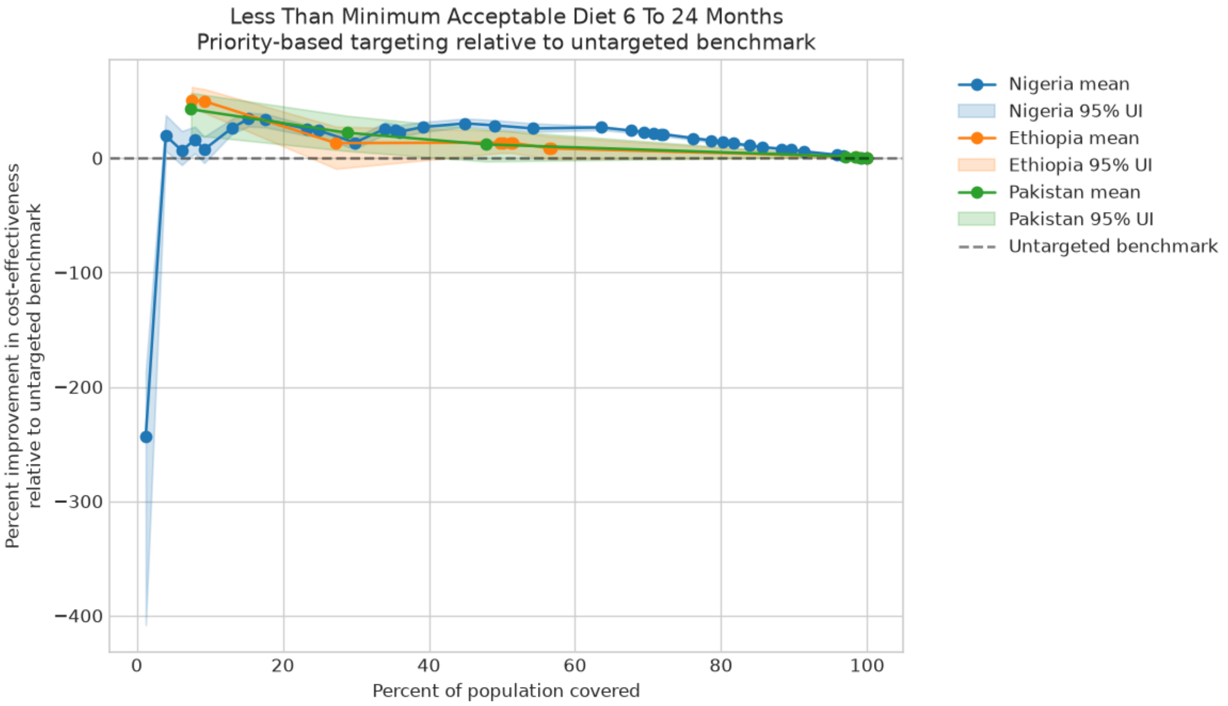


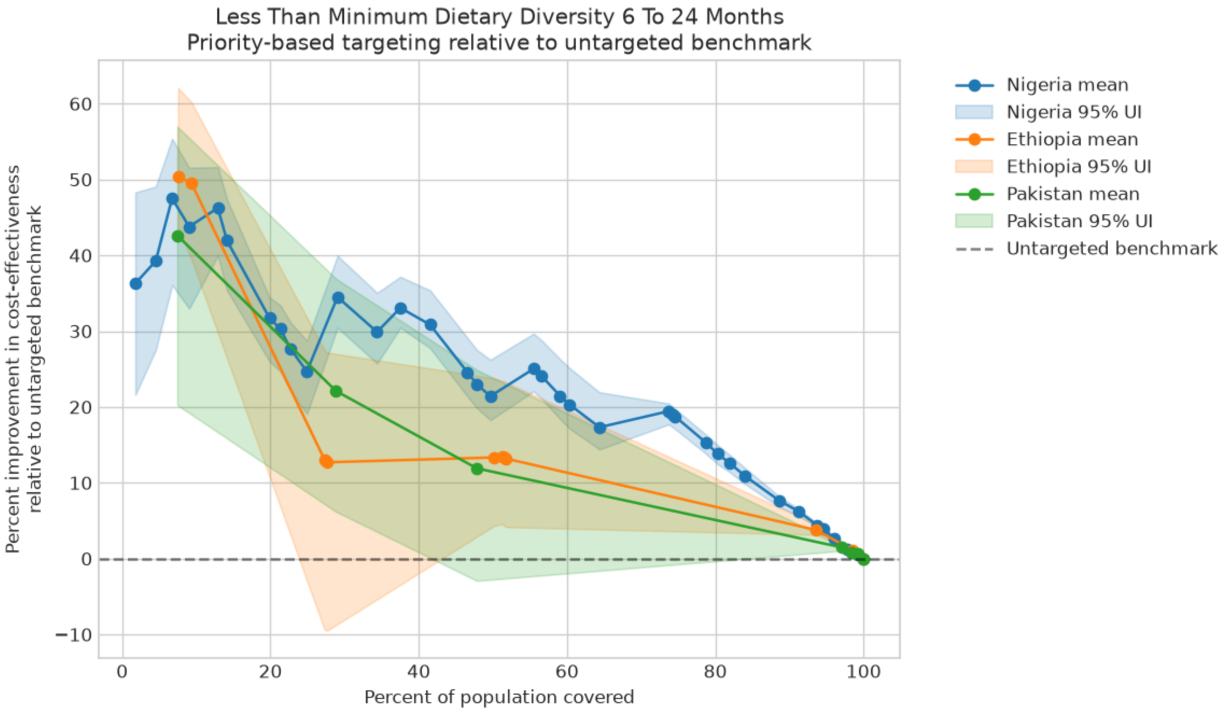

Figure A4-1: Priority-based targeting indicator cost-effectiveness relative to the untargeted benchmark (continued)

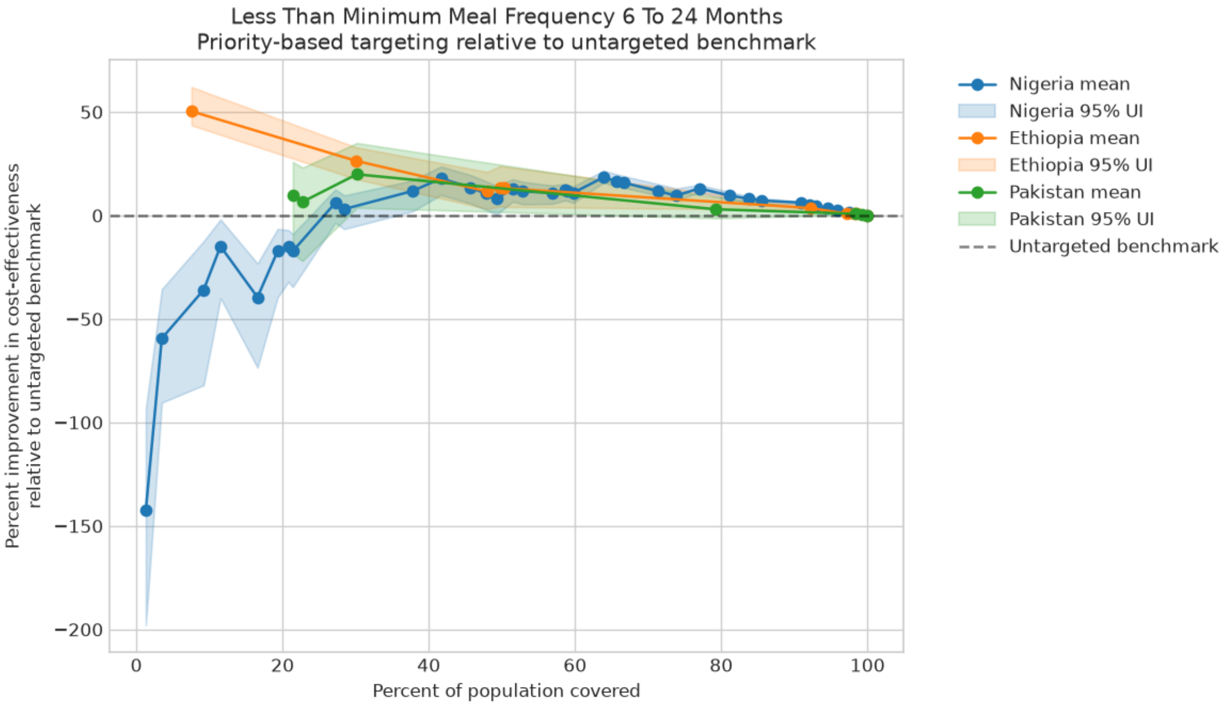


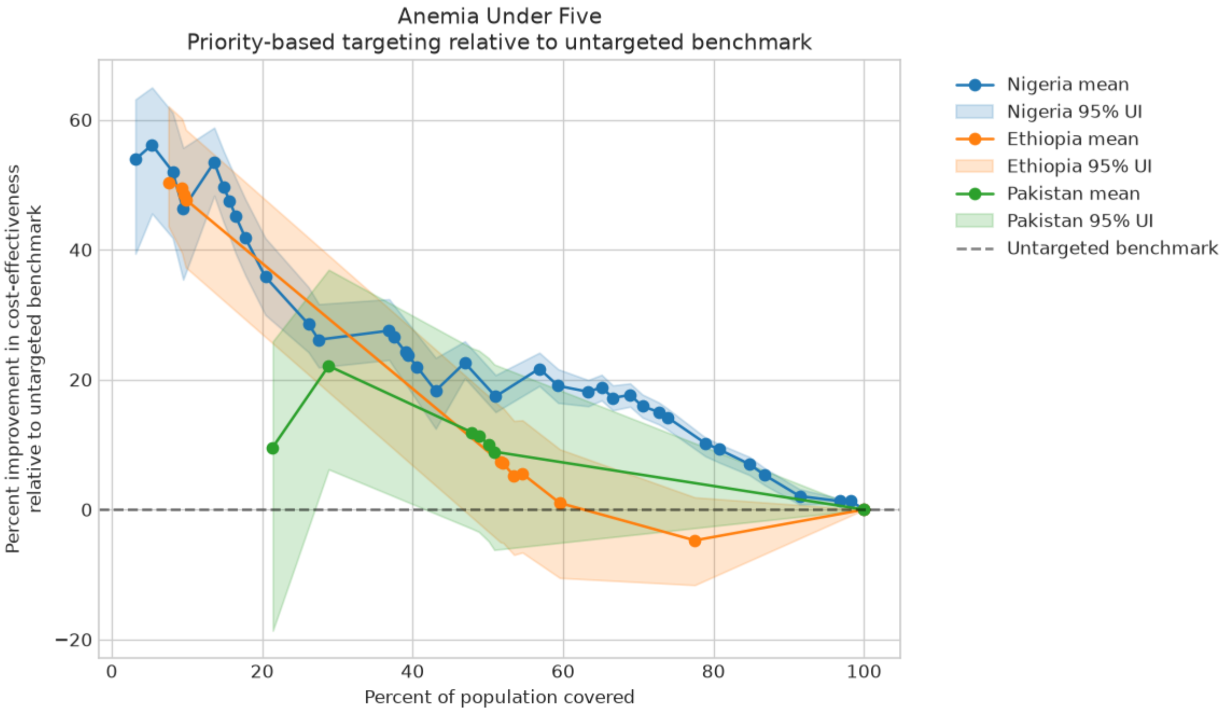

**Appendix 5: SQ-LNS effect modification results**

Table A5-1: Subnational results of SQ-LNS in Ethiopia with SQ-LNS effects modified by baseline wasting burden

| Location | Annual DALYs at baseline among the 6-59 month population | Annual DALYs averted | Incremental cost-effectiveness ratio | Total incremental cost | Incremental cost: SQ-LNS | Incremental cost: acute malnutrition treatment |
|---|---|---|---|---|---|---|
| | | | | | (USD / DALY averted) | (Millions of USD / year) |
| Addis Ababa | 28,400 (22,800, 35,200) | 672 (364, 1,280) | 2,530 (1,160, 4,030) | 1.44 (1.35, 1.52) | 1.46 (1.36, 1.51) | -0.0193 (-0.0432, 0.0125) |
| Afar | 156,000 (129,000, 195,000) | 8,010 (3,950, 13,100) | 218 (118, 406) | 1.54 (1.44, 1.66) | 1.71 (1.60, 1.78) | -0.167 (-0.237, -0.0892) |
| Amhara | 749,000 (636,000, 893,000) | 19,100 (9,980, 33,000) | 1,040 (513, 1,740) | 17.1 (15.6, 18.0) | 17.3 (16.2, 18.0) | -0.267 (-0.950, 0.484) |
| Benishangul-Gumuz | 86,900 (74,700, 106,000) | 3,540 (1,460, 5,160) | 332 (204, 684) | 1.02 (0.944, 1.08) | 1.08 (1.01, 1.12) | -0.0613 (-0.0960, -0.0358) |
| Dire Dawa | 16,400 (13,900, 18,900) | 640 (357, 883) | 498 (326, 784) | 0.292 (0.271, 0.311) | 0.318 (0.298, 0.331) | -0.0258 (-0.0382, -0.0156) |
| Gambella | 18,800 (15,000, 24,400) | 712 (247, 1,140) | 528 (271, 1,280) | 0.308 (0.283, 0.329) | 0.340 (0.319, 0.354) | -0.0319 (-0.0457, -0.0187) |
| Harari | 6,110 (4,730, 7,740) | 165 (74.2, 311) | 1,250 (559, 2,450) | 0.178 (0.165, 0.187) | 0.182 (0.171, 0.189) | -0.00455 (-0.00939, -0.000992) |
| Oromia | 2,130,000 (1,780,000, 2,520,000) | 45,300 (-6,740, 95,200) | 847 (-501, 1,720) | 39.6 (36.7, 43.4) | 40.2 (37.6, 41.7) | -0.546 (-1.78, 2.35) |
| Somali | 670,000 (567,000, 790,000) | 34,200 (16,500, 54,300) | 203 (119, 409) | 6.24 (5.71, 6.88) | 7.07 (6.62, 7.36) | -0.827 (-1.17, -0.471) |
| Southern Nations, Nationalities, and Peoples | 1,460,000 (1,220,000, 1,700,000) | 67,200 (34,700, 102,000) | 323 (198, 576) | 20.0 (18.4, 21.3) | 21.7 (20.4, 22.6) | -1.73 (-2.50, -1.23) |
| Tigray | 153,000 (130,000, 178,000) | 6,390 (2,830, 8,860) | 804 (519, 1,630) | 4.57 (4.30, 4.81) | 4.82 (4.50, 5.00) | -0.245 (-0.375, -0.149) |
| **National** | **5,480,000 (4,640,000, 6,440,000)** | **186,000 (103,000, 300,000)** | **536 (305, 910)** | **92.3 (85.7, 98.6)** | **96.2 (90.0, 99.9)** | **-3.92 (-5.91, -0.283)** |

DALY: Disability-adjusted life year; USD: 2021 United States Dollar; SQ-LNS: small-quantity lipid-based nutrient supplementation

Table A5-2: Subnational results of SQ-LNS in Nigeria with SQ-LNS effects modified by baseline wasting burden

| Location | Disability adjusted life years at baseline among the 6-59 month population | Disability adjusted life years averted | Incremental cost-effectiveness ratio | Total incremental cost (Millions of USD) | Incremental cost: SQ-LNS | Incremental cost: acute malnutrition treatment |
|---|---|---|---|---|---|---|
| | | | (USD) | (Millions of USD) | | |
| Abia | 459,000 (399,000, 517,000) | 10,400 (3,190, 16,300) | 723 (335, 1,760) | 5.40 (5.24, 5.63) | 5.54 (5.34, 5.72) | -0.134 (-0.316, -0.0106) |
| Adamawa | 630,000 (544,000, 692,000) | 15,200 (6,010, 24,200) | 375 (183, 766) | 4.51 (4.32, 4.71) | 4.57 (4.38, 4.76) | -0.0683 (-0.114, 0.0372) |
| Akwa Ibom | 145,000 (124,000, 172,000) | 3,380 (1,120, 5,620) | 932 (418, 2,140) | 2.34 (2.26, 2.44) | 2.39 (2.30, 2.47) | -0.0558 (-0.117, -0.0102) |
| Anambra | 411,000 (353,000, 455,000) | 7,640 (-1,700, 17,600) | 931 (-2,010, 3,580) | 7.50 (7.21, 7.70) | 7.60 (7.30, 7.86) | -0.0937 (-0.274, 0.0456) |
| Bauchi | 2,360,000 (2,010,000, 2,690,000) | 73,900 (32,700, 146,000) | 191 (79.3, 362) | 11.3 (10.8, 11.9) | 11.6 (11.1, 12.0) | -0.244 (-0.365, -0.0777) |
| Bayelsa | 58,600 (49,100, 67,800) | 1,350 (525, 2,300) | 818 (380, 1,760) | 0.869 (0.839, 0.893) | 0.880 (0.845, 0.905) | -0.0111 (-0.0158, -0.00556) |
| Benue | 952,000 (851,000, 1,030,000) | 19,800 (5,030, 32,800) | 903 (376, 2,520) | 12.1 (11.7, 12.7) | 12.3 (11.8, 12.7) | -0.118 (-0.315, 0.0646) |
| Borno | 500,000 (422,000, 560,000) | 31,800 (8,000, 41,600) | 279 (150, 845) | 6.22 (6.01, 6.37) | 6.67 (6.43, 6.90) | -0.453 (-0.575, -0.332) |
| Cross River | 136,000 (125,000, 154,000) | 2,800 (1,380, 4,560) | 1,020 (579, 1,890) | 2.52 (2.42, 2.59) | 2.56 (2.46, 2.64) | -0.0361 (-0.0567, 0.00127) |
| Delta | 314,000 (286,000, 338,000) | 6,700 (2,450, 16,600) | 1,070 (307, 1,930) | 4.74 (4.57, 4.89) | 4.83 (4.63, 5.00) | -0.0867 (-0.143, -0.00705) |
| Ebonyi | 322,000 (272,000, 359,000) | 9,630 (2,820, 20,000) | 621 (208, 1,690) | 4.08 (3.93, 4.19) | 4.18 (4.01, 4.29) | -0.0958 (-0.166, -0.0306) |
| Edo | 309,000 (263,000, 356,000) | 5,510 (1,290, 10,500) | 1,250 (381, 3,190) | 4.03 (3.87, 4.14) | 4.08 (3.92, 4.22) | -0.0462 (-0.102, 0.0851) |
| Ekiti | 120,000 (93,900, 139,000) | 2,370 (613, 3,880) | 1,180 (470, 3,040) | 1.82 (1.75, 1.87) | 1.83 (1.76, 1.88) | -0.0180 (-0.0363, 0.00352) |
| Enugu | 223,000 (198,000, 239,000) | 4,170 (1,400, 7,470) | 1,600 (705, 3,780) | 5.05 (4.86, 5.19) | 5.10 (4.90, 5.25) | -0.0533 (-0.0897, -0.0183) |
| FCT (Abuja) | 430,000 (348,000, 500,000) | 8,740 (2,760, 15,200) | 1,170 (486, 3,060) | 7.42 (7.17, 7.68) | 7.50 (7.18, 7.76) | -0.0770 (-0.148, -0.00777) |
| Gombe | 830,000 (692,000, 955,000) | 57,200 (22,500, 84,200) | 111 (59.6, 231) | 5.12 (4.96, 5.32) | 5.34 (5.13, 5.49) | -0.225 (-0.329, -0.155) |
| Imo | 633,000 (546,000, 729,000) | 12,100 (4,040, 21,900) | 1,390 (559, 3,060) | 12.2 (11.8, 12.6) | 12.4 (11.9, 12.8) | -0.133 (-0.315, 0.0532) |
| Jigawa | 1,910,000 (1,600,000, 2,160,000) | 131,000 (47,000, 188,000) | 85.8 (46.5, 197) | 8.92 (8.67, 9.20) | 9.45 (9.10, 9.73) | -0.526 (-0.743, -0.396) |
| Kaduna | 1,570,000 (1,380,000, 1,730,000) | 92,500 (29,500, 146,000) | 220 (103, 519) | 14.9 (14.4, 15.5) | 15.8 (15.1, 16.3) | -0.818 (-0.981, -0.588) |
| Kano | 4,870,000 (4,070,000, 5,490,000) | 281,000 (65,500, 414,000) | 143 (63.7, 425) | 26.3 (25.5, 27.4) | 27.9 (26.7, 28.9) | -1.54 (-2.16, -1.19) |
| Katsina | 2,890,000 (2,530,000, 3,210,000) | 214,000 (73,400, 313,000) | 94.6 (52.6, 230) | 16.0 (15.6, 16.6) | 17.1 (16.5, 17.8) | -1.09 (-1.71, -0.830) |
| Kebbi | 1,320,000 (1,110,000, 1,440,000) | 96,400 (32,900, 155,000) | 104 (49.3, 238) | 7.55 (7.29, 7.76) | 8.11 (7.82, 8.34) | -0.564 (-0.779, -0.455) |
| Kogi | 196,000 (180,000, 213,000) | 4,150 (1,380, 7,270) | 1,120 (436, 2,370) | 3.23 (3.11, 3.40) | 3.31 (3.19, 3.41) | -0.0732 (-0.182, 0.0406) |
| Kwara | 172,000 (144,000, 194,000) | 3,440 (565, 5,860) | 2,080 (602, 8,520) | 3.62 (3.48, 3.74) | 3.67 (3.53, 3.79) | -0.0517 (-0.0895, -0.00851) |
| Lagos | 236,000 (199,000, 254,000) | 4,940 (836, 9,670) | 2,140 (543, 8,800) | 5.20 (5.04, 5.38) | 5.33 (5.12, 5.50) | -0.131 (-0.236, -0.0377) |
| Nasarawa | 555,000 (434,000, 628,000) | 14,800 (2,570, 26,000) | 1,030 (296, 3,700) | 7.81 (7.44, 8.10) | 7.88 (7.58, 8.10) | -0.0736 (-0.162, 0.0942) |
| Niger | 1,690,000 (1,470,000, 1,920,000) | 41,300 (9,400, 74,000) | 676 (227, 1,880) | 16.7 (16.1, 17.5) | 16.9 (16.3, 17.6) | -0.219 (-0.503, 0.270) |
| Ogun | 784,000 (675,000, 877,000) | 21,100 (5,180, 34,200) | 981 (388, 2,930) | 13.6 (13.0, 14.3) | 13.8 (13.3, 14.3) | -0.262 (-0.422, 0.114) |
| Ondo | 264,000 (230,000, 299,000) | 6,830 (2,100, 13,200) | 797 (293, 1,850) | 3.85 (3.73, 3.98) | 3.94 (3.78, 4.08) | -0.0856 (-0.135, -0.0333) |
| Osun | 177,000 (157,000, 194,000) | 3,200 (989, 5,590) | 1,970 (802, 4,800) | 4.49 (4.29, 4.59) | 4.55 (4.37, 4.69) | -0.0585 (-0.111, 0.102) |
| Oyo | 309,000 (262,000, 348,000) | 6,490 (1,390, 12,100) | 1,620 (522, 5,780) | 6.30 (6.04, 6.50) | 6.38 (6.13, 6.58) | -0.0811 (-0.124, -0.0116) |
| Plateau | 936,000 (820,000, 1,040,000) | 26,200 (5,800, 42,900) | 952 (366, 2,720) | 15.5 (14.9, 16.0) | 15.7 (15.1, 16.2) | -0.213 (-0.349, -0.0249) |
| Rivers | 285,000 (230,000, 326,000) | 5,790 (1,180, 12,000) | 1,150 (321, 3,590) | 3.86 (3.69, 3.97) | 3.91 (3.76, 4.02) | -0.0441 (-0.0706, -0.0160) |
| Sokoto | 2,770,000 (2,450,000, 3,060,000) | 201,000 (90,400, 284,000) | 64.5 (40.1, 126) | 11.1 (10.6, 11.7) | 12.1 (11.6, 12.6) | -1.00 (-1.40, -0.725) |
| Taraba | 1,490,000 (1,190,000, 1,730,000) | 38,500 (16,600, 68,500) | 396 (175, 723) | 11.9 (11.5, 12.2) | 12.1 (11.6, 12.4) | -0.181 (-0.269, -0.0517) |
| Yobe | 815,000 (689,000, 926,000) | 56,200 (24,200, 90,500) | 139 (70.3, 266) | 6.35 (6.20, 6.53) | 6.83 (6.58, 7.05) | -0.477 (-0.560, -0.355) |
| Zamfara | 1,640,000 (1,390,000, 1,810,000) | 109,000 (35,500, 158,000) | 74.9 (39.1, 184) | 6.28 (6.06, 6.50) | 6.65 (6.41, 6.85) | -0.374 (-0.513, -0.305) |
| **National** | **33,700,000 (30,600,000, 36,300,000)** | **1,630,000 (598,000, 2,280,000)** | **225 (128, 492)** | **291 (281, 301)** | **301 (289, 311)** | **-9.81 (-13.2, -7.29)** |

DALY: Disability-adjusted life year; USD: 2021 United States Dollar; SQ-LNS: small-quantity lipid-based nutrient supplementation

Table A5-3: Subnational results of SQ-LNS in Pakistan with SQ-LNS effects modified by baseline wasting burden

| Location | Disability adjusted life years at baseline among the 6-59 month population | Disability adjusted life years averted | Incremental cost-effectiveness ratio (USD) | Total incremental cost (Millions of USD) | Incremental cost: SQ-LNS (Millions of USD) | Incremental cost: acute malnutrition treatment (Millions of USD) |
|---|---|---|---|---|---|---|
| Azad Jammu & Kashmir | 117,000 (87,200, 165,000) | 2,290 (805, 3,600) | 1,860 (907, 5,700) | 3.29 (3.14, 3.43) | 3.29 (3.14, 3.43) | -0.00221 (-0.00507, 0.000966) |
| Balochistan | 967,000 (769,000, 1,280,000) | 73,800 (44,600, 112,000) | 259 (155, 400) | 17.6 (16.8, 18.4) | 17.8 (17.0, 18.5) | -0.207 (-0.293, -0.136) |
| Gilgit-Baltistan | 78,100 (62,200, 106,000) | 2,150 (879, 3,640) | 1,240 (601, 2,560) | 2.26 (2.16, 2.35) | 2.26 (2.17, 2.36) | -0.00395 (-0.00901, -0.000960) |
| Islamabad Capital Territory | 17,800 (14,100, 21,400) | 566 (317, 930) | 3,650 (2,080, 5,730) | 1.86 (1.78, 1.94) | 1.87 (1.79, 1.94) | -0.00269 (-0.00674, 0.00212) |
| Khyber Pakhtunkhwa | 1,450,000 (1,160,000, 1,960,000) | 90,400 (55,400, 132,000) | 545 (337, 846) | 45.4 (43.4, 47.3) | 45.7 (43.8, 47.6) | -0.340 (-0.460, -0.223) |
| Punjab | 4,260,000 (3,490,000, 5,370,000) | 121,000 (57,400, 191,000) | 1,090 (617, 2,140) | 119 (113, 124) | 119 (114, 124) | -0.338 (-0.739, 0.0463) |
| Sindh | 1,820,000 (1,390,000, 2,460,000) | 128,000 (70,400, 189,000) | 436 (268, 736) | 51.5 (49.3, 53.6) | 51.9 (49.7, 54.1) | -0.450 (-0.675, -0.272) |
| **National** | **8,720,000 (7,260,000, 11,200,000)** | **418,000 (268,000, 542,000)** | **609 (439, 908)** | **241 (230, 251)** | **242 (231, 252)** | **-1.34 (-1.70, -0.798)** |

DALY: Disability-adjusted life year; USD: 2021 United States Dollar; SQ-LNS: small-quantity lipid-based nutrient supplementation

Table A5-4: Threshold-Based Targeting Algorithm Results, Effect Modification Sensitivity Analysis

| Threshold | Ethiopia | | | | Nigeria | | | | Pakistan | | | |
|---|---|---|---|---|---|---|---|---|---|---|---|---|
| | Coverage | Annual DALYs Averted (thousands) | Annual Incremental Cost (millions of USD) | ICER | Coverage | Annual DALYs Averted (thousands) | Annual Incremental Cost (millions of USD) | ICER | Coverage | Annual DALYs Averted (thousands) | Annual Incremental Cost (millions of USD) | ICER |
| High anemia (>40%) | 100 | 186 (103, 300) | 92.3 (85.7, 98.6) | 536 (305, 910) | 100 | 1,631 (598, 2,280) | 291 (281, 301) | 225 (128, 492) | 100 | 418 (268, 542) | 241 (230, 251) | 609 (439, 908) |
| High stunting (>20%) | 98.6 | 185 (102, 300) | 90.8 (84.4, 97.1) | 530 (301, 899) | 79.8 | 1,558 (574, 2,157) | 230 (223, 238) | 186 (107, 408) | 98.7 | 416 (266, 540) | 237 (227, 247) | 604 (434, 902) |
| High wasting (>10%) | 38.6 | 121 (60.7, 175) | 34.0 (31.5, 36.3) | 305 (197, 567) | 39 | 1,271 (461, 1,689) | 109 (106, 113) | 108 (64.0, 243) | 47.8 | 292 (177, 397) | 114 (109, 119) | 418 (283, 651) |
| High wasting + stunting + anemia | 38.6 | 121 (60.7, 175) | 34.0 (31.5, 36.3) | 305 (197, 567) | 39 | 1,271 (461, 1,689) | 109 (106, 113) | 108 (64.0, 243) | 47.8 | 292 (177, 397) | 114 (109, 119) | 418 (283, 651) |
| GHI > 20 (serious) | 98.6 | 185 (102, 300) | 90.8 (84.4, 97.1) | 530 (301, 899) | 79.6 | 1,566 (578, 2,173) | 229 (222, 238) | 185 (106, 405) | 97.9 | 415 (265, 539) | 235 (225, 245) | 600 (432, 897) |
| GHI > 35 (alarming) | 1.8 | 8.01 (3.95, 13.1) | 1.54 (1.44, 1.66) | 218 (118, 406) | 40.7 | 1,313 (483, 1,785) | 114 (111, 118) | 109 (64.4, 244) | 28.8 | 202 (115, 287) | 69.1 (66.1, 72.0) | 368 (236, 605) |

DALY: Disability-adjusted life year, ICER: Incremental cost-effectiveness ratio, GHI: Global Hunger Index Score, USD: 2021 United States Dollars

Figure A5-1: Priority-based targeting indicator cost-effectiveness relative to the untargeted benchmark with SQ-LNS effect modification

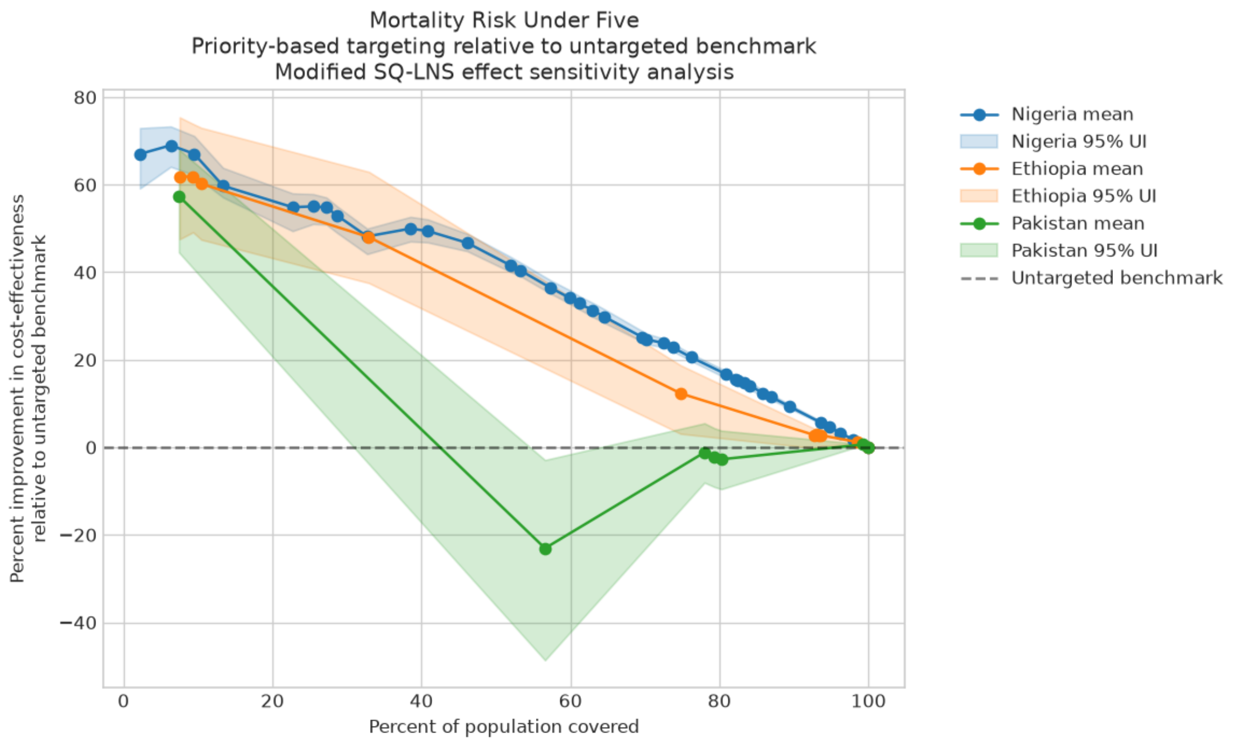


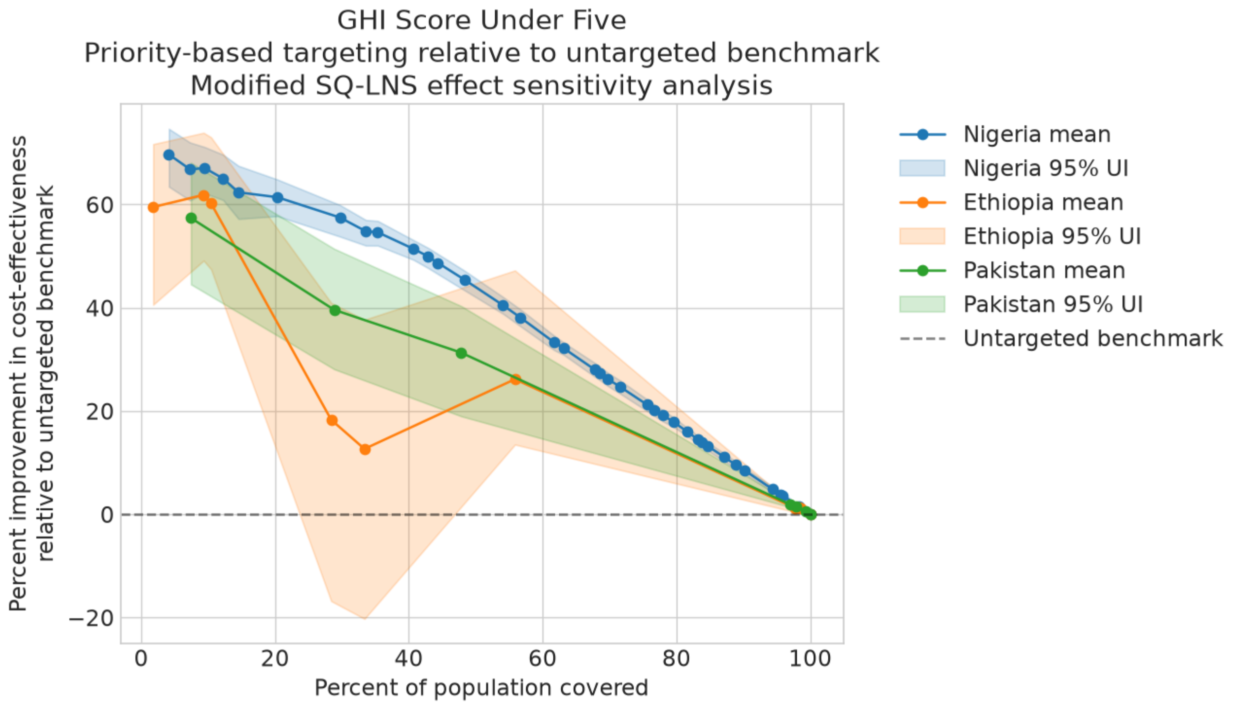

Figure A5-1: Priority-based targeting indicator cost-effectiveness relative to the untargeted benchmark with SQ-LNS effect modification (continued)

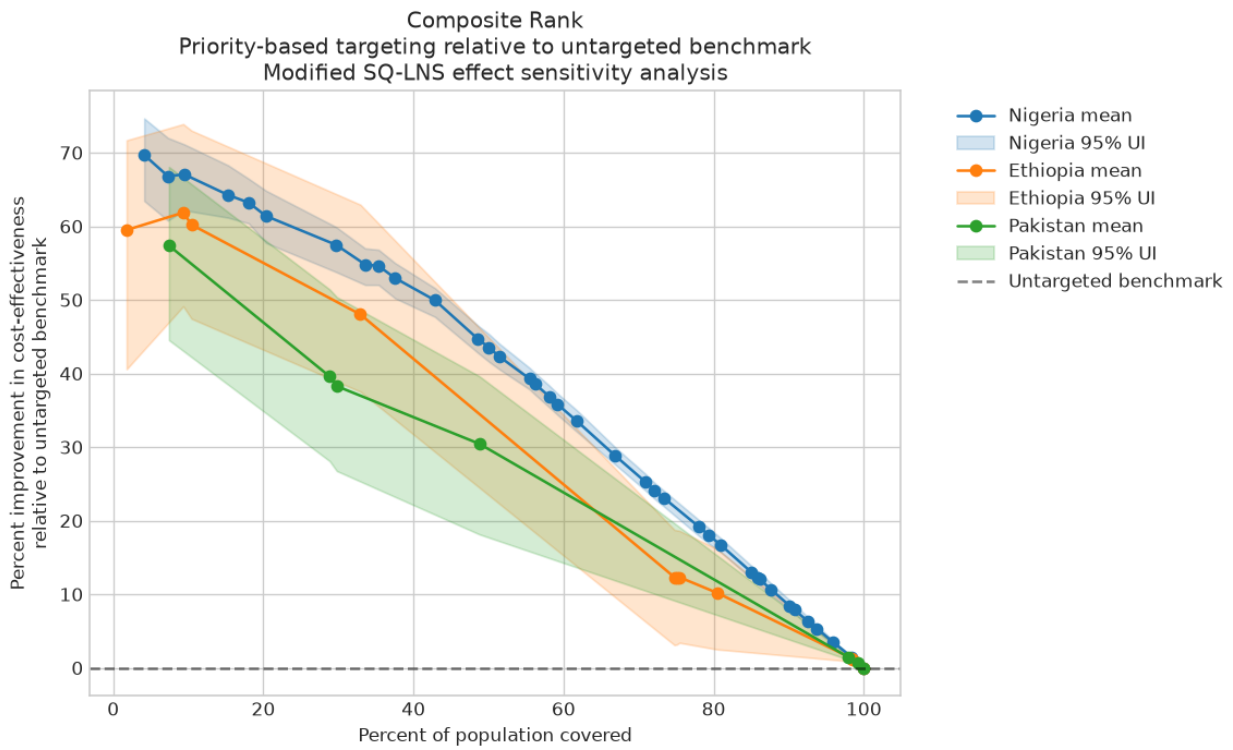


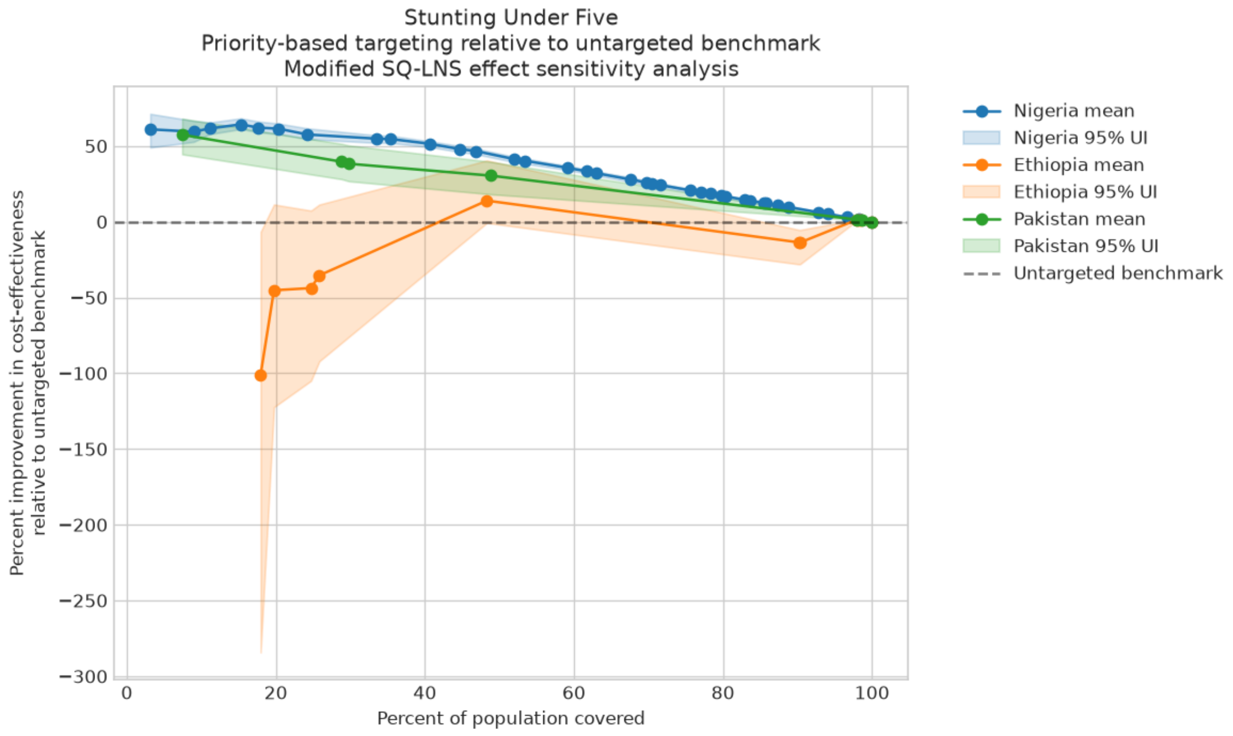

Figure A5-1: Priority-based targeting indicator cost-effectiveness relative to the untargeted benchmark with SQ-LNS effect modification (continued)

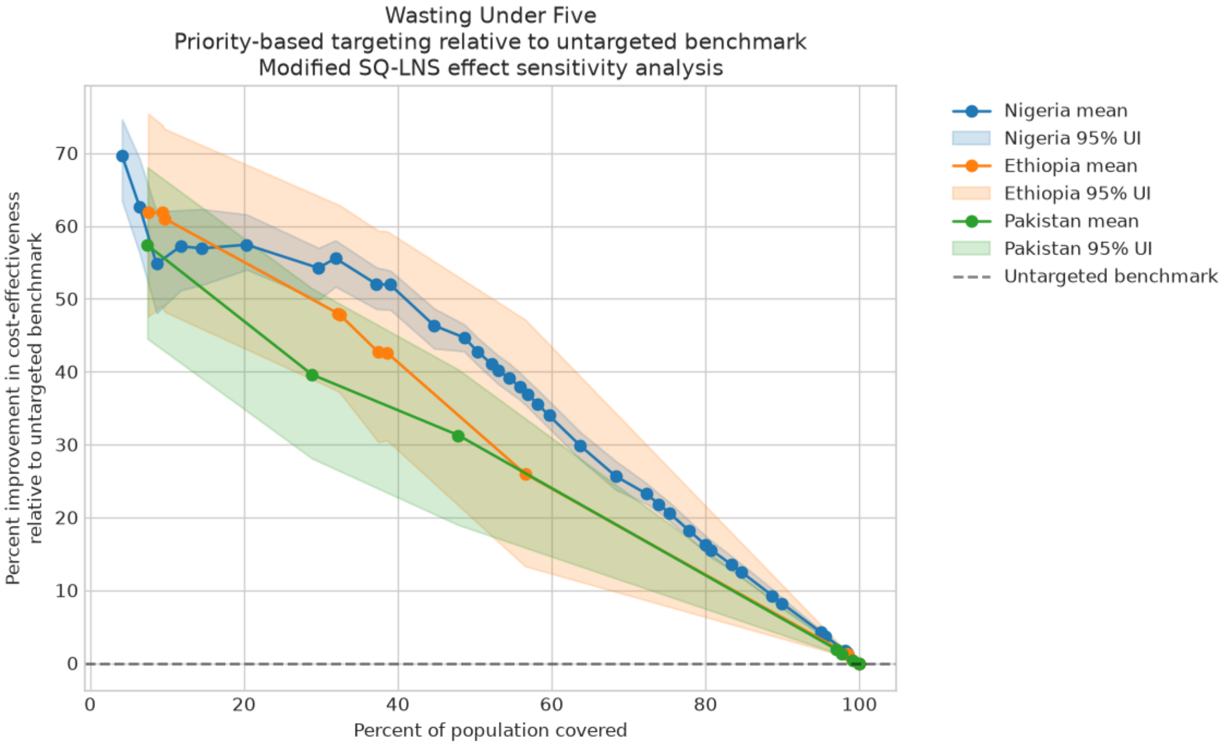


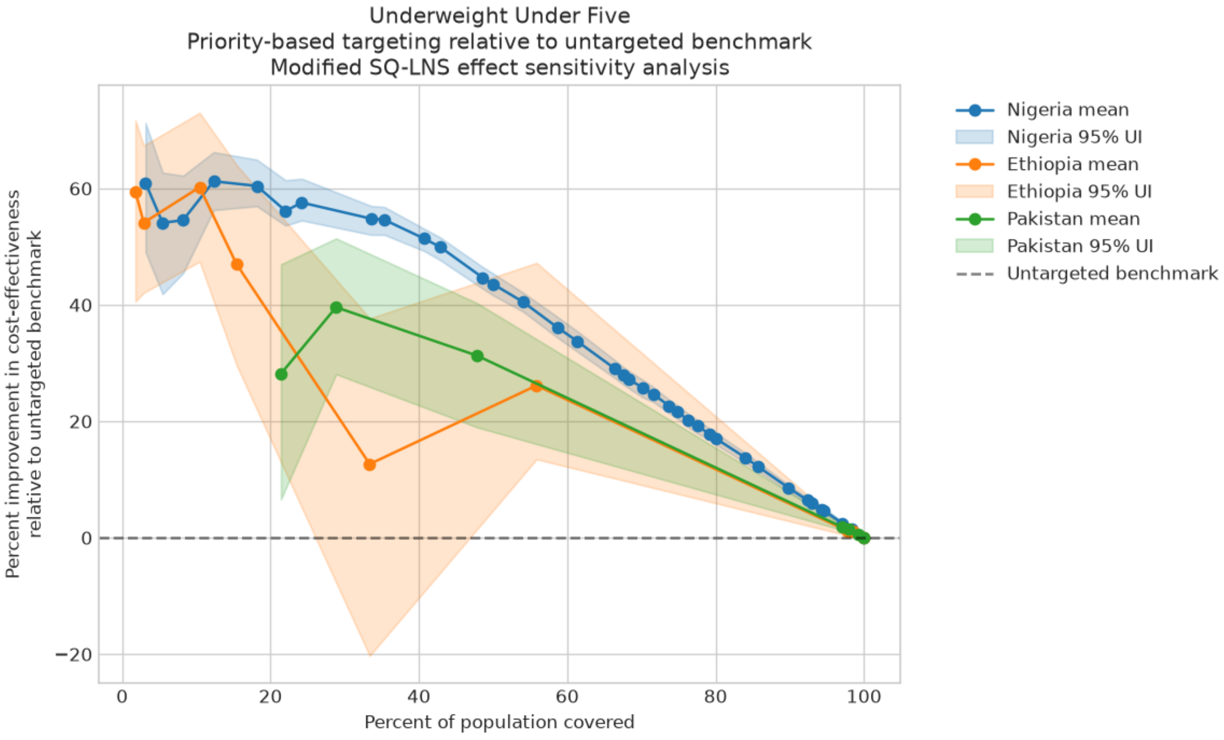

Figure A5-1: Priority-based targeting indicator cost-effectiveness relative to the untargeted benchmark with SQ-LNS effect modification (continued)

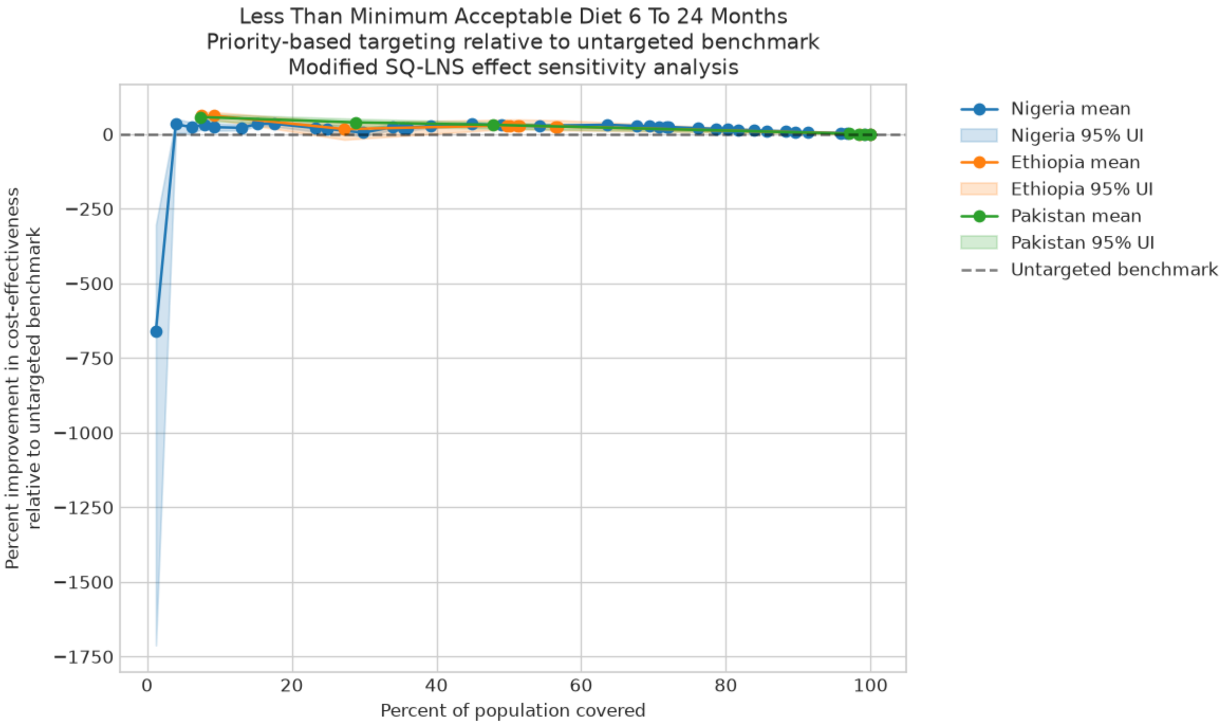


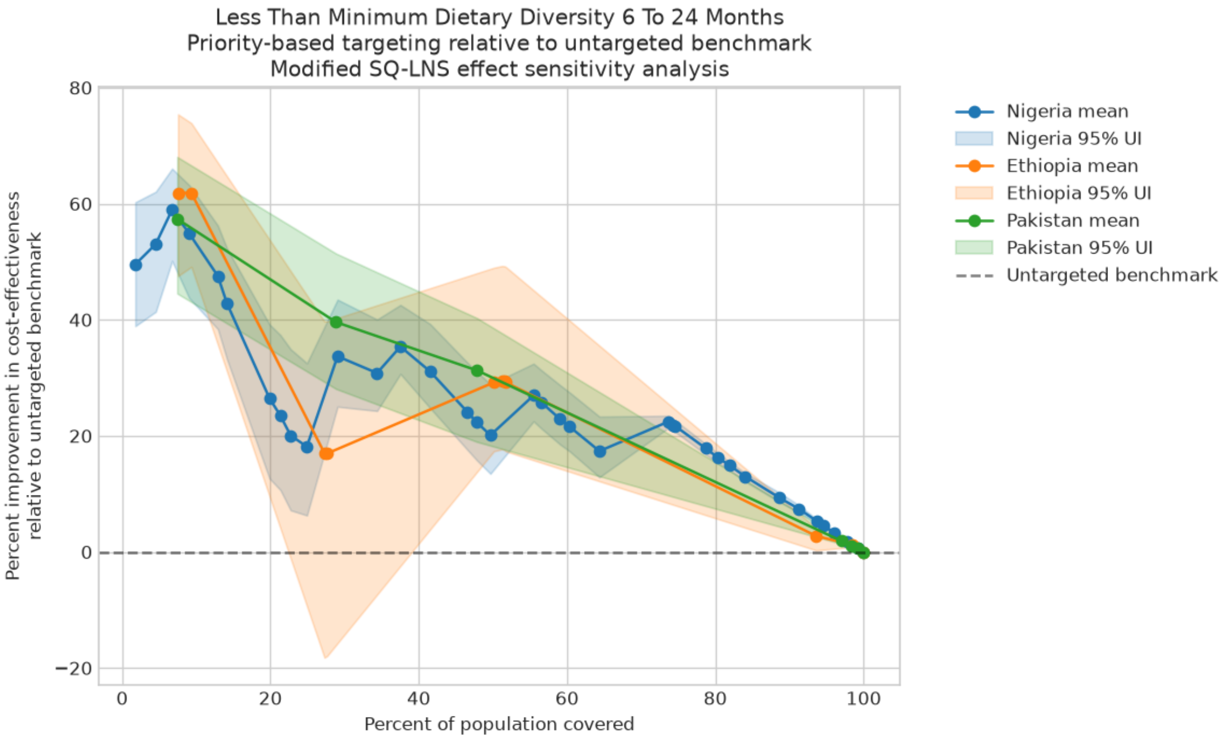

Figure A5-1: Priority-based targeting indicator cost-effectiveness relative to the untargeted benchmark with SQ-LNS effect modification (continued)

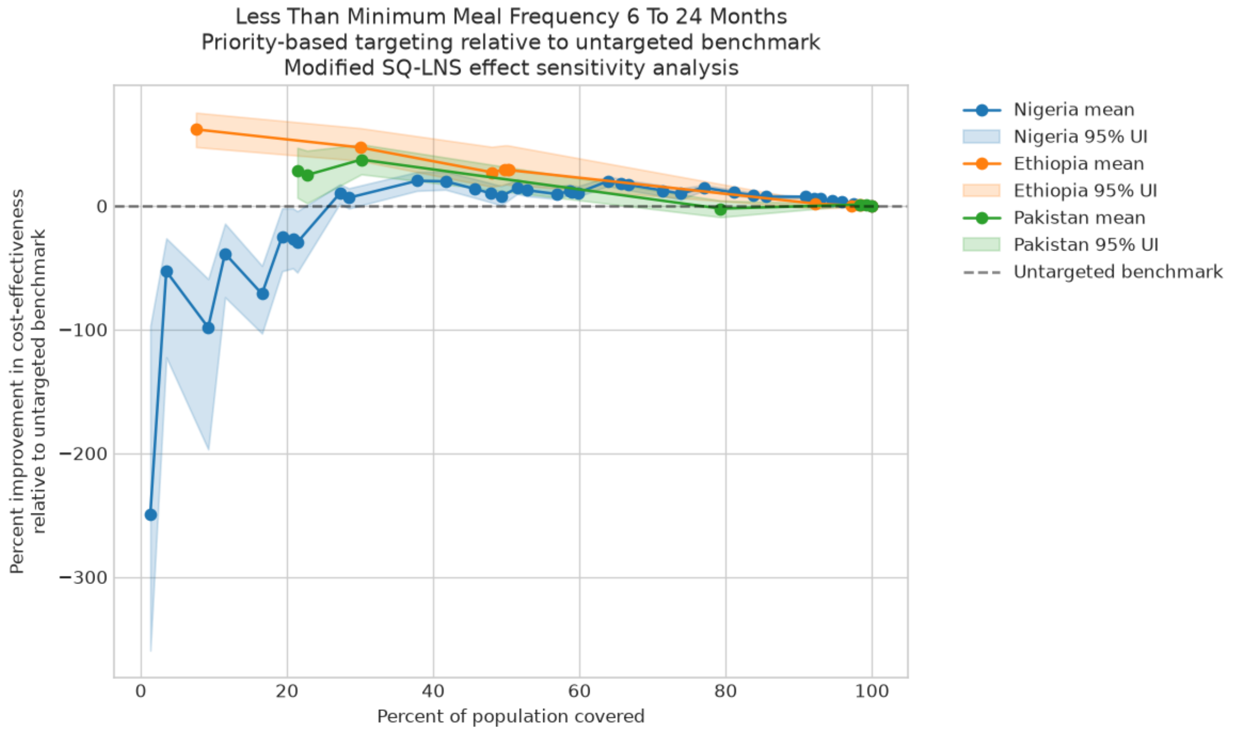


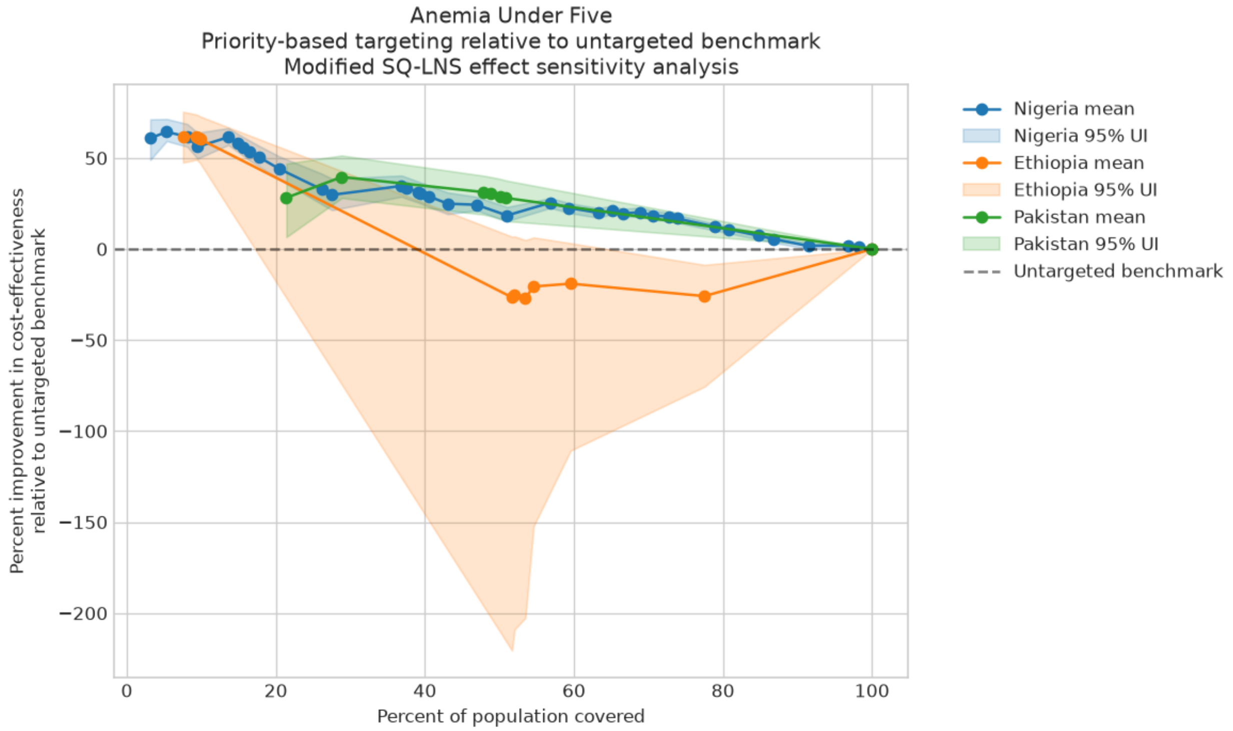
